# Assessing spoken discourse in aphasia using multimodal artificial intelligence

**DOI:** 10.64898/2026.09.27.26364108

**Authors:** M. J. Marte, S. Lee, S. Wang, K. Goldin, M. Varkanitsa, J.M. Girard, R. Brito, X. Wei, J.T. Baker, Y. Tie, S. Kiran, E. Liebenthal

## Abstract

Discourse analysis can reliably predict real-world communicative success, yet is rarely implemented clinically, due to challenges in manually generating stimulus-specific main concept inventories (MCIs) and scoring patient narratives. We evaluated whether automatically derived macrolinguistic (main concept, sentiment) and microlinguistic (fluency, lexical, syntactic) features extracted from movie clip narrations can augment the clinical assessment of discourse in aphasia. The MCIs generated directly from each clip’s video, audio and subtitles using a vision-language model approximated the main concepts healthy controls produced. Group classification (control: n=51; aphasia: n=54) based on the discourse features was excellent (AUC=0.95, 95% CI [0.90, 0.99]), with macrolinguistic features showing the largest group differences (main concept completeness: d=-1.84, 95% CI [−2.30, −1.38]) and strongest associations with aphasia severity (semantic distance to inventory: r=-0.79, 95% CI [−0.87, −0.66]). Findings support the feasibility and scalability of automated criterion-based discourse assessment using ecological stimuli, and highlight its potential for clinical implementation.

## Background

More than two million individuals in the United States currently suffer from post-stroke aphasia, with discourse production and comprehension impairments representing a primary barrier to functional communication and community reintegration^1,2^. Despite evidence that discourse measures predict real-world communicative success better than standardized single-word tasks^3–5^, discourse analysis remains largely confined to research settings^6,7^. A primary challenge to clinical implementation is that most discourse measures lack sufficient psychometric definition to be used as diagnostic or outcome measures^8^. Another limitation of standard aphasia batteries is the use of static, decontextualized (and often outdated) stimuli to elicit discourse. The single-picture “Cookie Theft Scene” used in the Boston Diagnostic Aphasia Examination, and the “Picnic Scene” used in the Western Aphasia Battery-Revised^9,10^, fail to engage temporally unfolding aspects of narrative comprehension and production. Thus, these tests poorly approximate the dynamic, multimodal demands of storytelling and conversation associated with real-world communication^11^. Additional validity challenges have been identified, for example, Kong^12^ reported that Cantonese speakers systematically misidentified the Cookie Theft kitchen setting and failed to recognize the cookie-stealing theme altogether.

Main concept analysis (MCA) is a criterion-based approach to evaluating discourse informativeness. MCA was shown to be sensitive to aphasia severity^13^ and treatment-induced changes^14^. MCA returns a quantitative estimate of the degree to which individual speakers produce essential semantic propositions that capture the “gist” of a story or event, relative to a main concept inventory developed from healthy control performance. The MCA approach is applicable to any stimulus, and could in principle be used to develop culturally appropriate and ecologically valid assessment materials, while providing objective, quantifiable metrics of discourse performance^4,15,16^. Nevertheless, prohibitive barriers to widespread clinical adoption of MCA remain, including the paucity of stimulus materials with associated normative reference inventories, and metrics of performance in clinical populations. Generating gold-standard normative main concept inventories manually is an arduous process requiring iterative analysis of a sizable set of healthy control discourse samples to identify the essential concepts for each stimulus. The main concepts have variably been defined as those produced orally by at least 33% of a normative sample^4^, or those written by at least 70% of a clinician panel and produced by at least 70% of a normative sample^15^, exemplifying the challenge of developing objective normative inventories. Clinical implementation also requires clinicians to undergo specialized training to achieve high inter-rater reliability in manual scoring of patient samples against the normative inventory^17^. Currently, main concept inventories exist only for a handful of stimuli, e.g., the Cookie Theft picture^16^, the Cinderella story^17^, and the Peanut Butter and Jelly procedure^4^, and their use in clinical practice is largely limited to manual evaluation.

Recent computational advances help address these barriers. Large language models (LLMs) can process discourse to generate structured outputs for automated MCA, and preliminary work has yielded promising results: Kurland et al.^18^ showed that different LLMs (GPT-4, GPT-4o, and Llama-3-70B) could consistently rate participant retellings of eight short audiovisual stimuli from the Brief Assessment of Transactional Success (BATS) against human-developed main concept inventories (MCIs), and yield strong agreement with expert human raters (r = 0.89–0.90). Gupta et al.^19^ further used an LLM prompt-ensemble method to automatically recover most of the main concepts from the text transcripts of the same BATS stimuli. However, in these initial studies, only categorical scores representing the presence or absence of a set of concepts in a narrative sample were computed. Continuous scores, representing the semantic distance from the narrative centroid, are more informative and better suited for predictive modeling, longitudinal monitoring, and integration with other computational discourse measures.

Lee et al.^20^ recently developed an automated embedding-based MCA pipeline for Cinderella story retellings relative to an existing human-developed inventory, paving the way to computing a continuous distance from a narration to an inventory centroid. Furthermore, computational methods have not yet been applied to generate main concept inventories directly from a multimodal stimulus. Vision-language models (VLMs), which are multimodal AI systems that integrate visual and text information in a single model, now enable machines to appraise components of multimodal stimuli conveyed not only by their verbal but also by their nonverbal content.

The overall goal of this study was to evaluate whether automated analysis of patient narrations of naturalistic movie clips can serve as a clinically valid, scalable assessment of the extent of discourse impairment in persons with aphasia (PWA). Specifically, we evaluated: (1) The quality of VLM-generated main MCIs relative to a reference of main concepts produced by healthy controls (HC). (2) The value of automatically derived macrolinguistic (i.e., main concept, sentiment) and microlinguistic (i.e., speech fluency, lexical, and syntactic) features^20,21^ extracted from the movie clip narrations for classification of PWA versus HC, and prediction of aphasia severity. We hypothesized that both macrolinguistic and microlinguistic automatic features would discriminate between PWA and HC, consistent with prior evidence that impairments in aphasia extend beyond speech fluency and syntax, to narrative informativeness and coherence^13,16^. A corroboration of this hypothesis would support the validity of the automatic macrolinguistic and microlinguistic features. Furthermore, we hypothesized that main concept features representing the extent of deviation from normal narrative structure (i.e., main concept semantic distance from the inventory) would provide complementary power for predicting aphasia severity beyond categorical main concept (i.e., main concept completeness) and microlinguistic features alone.

## Results

### Validation of VLM-generated main concept inventories

A total of 54 concepts for the eight clips were found in at least three of the five runs of VLM inventory generation. Of those, 15 concepts were produced by fewer than 20% of HC, and were therefore excluded from the inventories. The remaining 39 concepts constituted the final inventories against which individual concept production was measured. The final inventory concepts were produced by HC at a median rate of 40% (Supplementary Table S1); mean production per clip ranged from 29.6% (concept range 22.5-40.0%) for “No Country for Old Men” to 62.4% (40.5-88.1%) for “Catch Me If You Can” (Supplementary Table S2). The number of inventory concepts per clip ranged 4-6 (mean = 4.9, SD = 0.8), and these concepts were proposed by 4.5 of the 5 VLM runs on average (27 of 39 by all five; Figure 1A).

**Figure 1.**
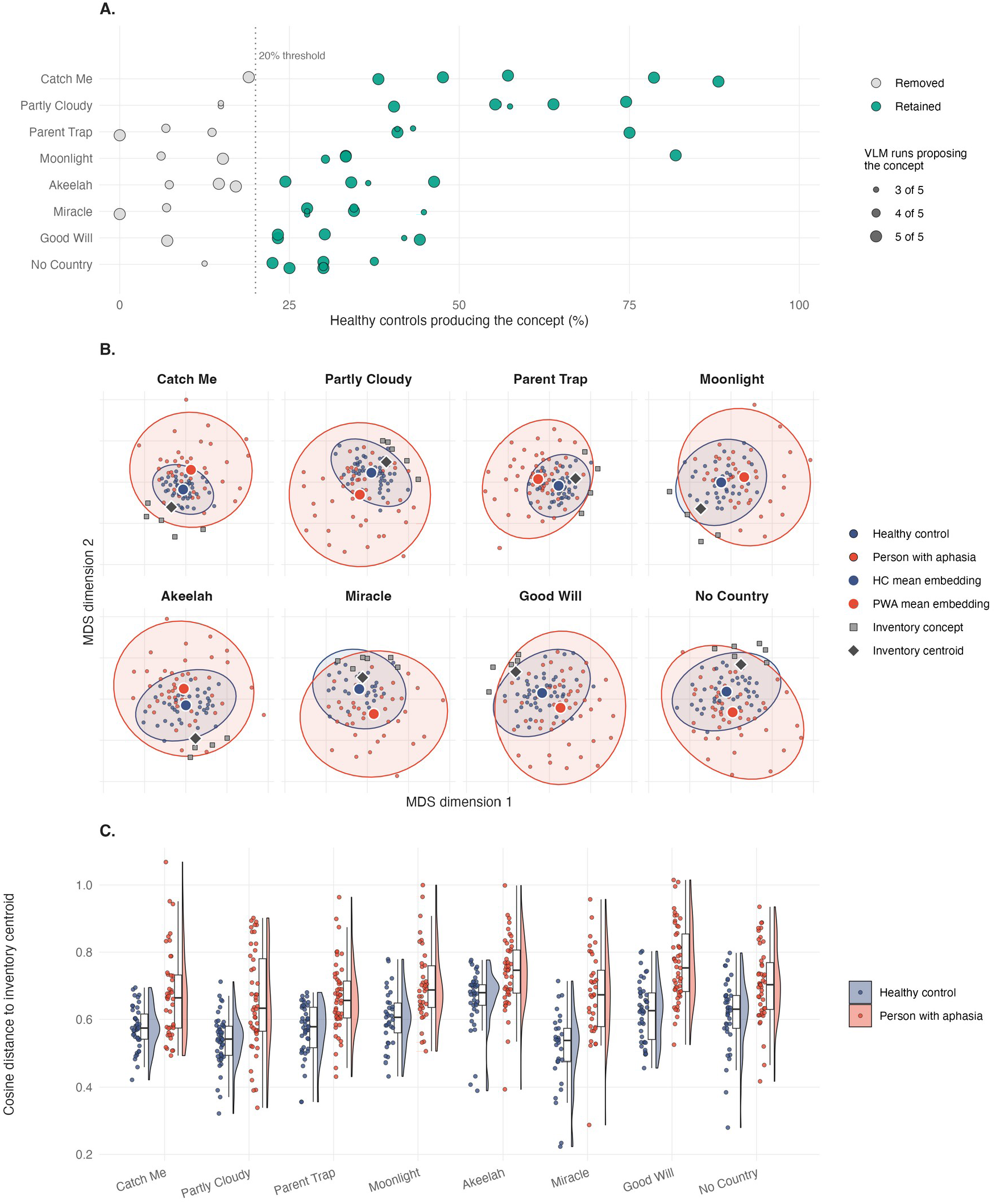
Main concept inventory construction and validation. (A) Each point on the graph represents one of the 54 concepts proposed by at least three of the five independent VLM runs, with point size indicating the number of runs that proposed the concept.

In the embedding space (projected for each clip in Figure 1B), the utterances nearest each clip’s inventory centroid were predominantly ones produced by HC. Across clips, HC contributed 72.3% of the top 5% of utterances closest to the centroid, against a base rate of 51.8% of all 5,054 utterances. Permuting participant group labels within clip placed the expected HC share at 48.5% (SD 4.9; z = 4.89, p < .001).

Seven of the inventory concepts were grounded strictly in audiovisual content (i.e., did not rely on a corresponding subtitle quote), including the 5 concepts of the dialogue-free animated clip, and 2 concepts from 2 other clips with dialogue. The audiovisual concepts were produced by HC at a mean rate of 53% (range 34-75%), within the same production range as the 32 concepts grounded in the subtitles (mean 41%, range 23-88%; Supplementary Table S1).

Points are placed on the x-axis by the percentage of HC who produced the concept. The dotted line indicates the 20% HC production rate threshold. The 39 concepts to its right (green) were retained in the final inventory, and the 15 concepts to its left (grey) were excluded. (B) Each inventory concept (grey squares) and the inventory centroid (grey diamond), and the group mean embeddings of HC (large blue circles) and PWA (large red circles) are presented for each clip in a two-dimensional metric multidimensional scaling of the embedding space. The small points represent the individual participants, each placed by the mean embedding of their utterances for that clip, colored by group (HC = blue; PWA = red); the shaded ellipses are 95% normal-probability ellipses per group. (C) Each point in the violin plots represents the cosine distance between a participant’s narration and the inventory centroid, by clip and group (HC=blue; PWA=red). Clips are ordered in all panels by mean HC production of their concepts, highest first.

### Group Comparisons on automatic macrolinguistic and microlinguistic features

The PWA and HC groups were compared on 4 macrolinguistic (main concept completeness, main concept distance to centroid, main concept semantic coherence, and sentiment) and 15 microlinguistic independent features covering multiple aspects of discourse processing (see descriptive statistics and group differences in Table 1). Ten of the 19 features differed between the groups after FDR correction (Table 1), with the macrolinguistic features showing overall larger group differences than the microlinguistic features (mean |d| = 1.05 versus 0.50; difference 0.55, 95% CI [0.24, 0.81]; exact permutation p = .033). Specifically, PWA relative to HC produced fewer unique main concepts (19% versus 46% of the inventory on average, respectively, d = −1.84, p < .001), and their utterances were semantically farther from the inventory narrative centroid (d = 1.41, p < .001, range: d = 0.82–1.38; Figure 1C, Supplementary Table S3). The sentiment ratings of the PWA movie narrations were more negative than the HC movie narrations (d = −0.87, p = .003). However, the local semantic coherence between consecutive utterances did not differ between groups (d = −0.07, p = .88). The microlinguistic features revealed moderate to large group differences across multiple domains. PWA exhibited higher disfluency rates (d = 1.07, p < .001) and produced fewer total words (d = −0.84, p < .001) than HC. Lexical diversity measures were higher in HC, including Honoré’s statistic (d = −0.84, p < .001) and word frequency range (d = −0.78, p < .001).

**Table 1.**
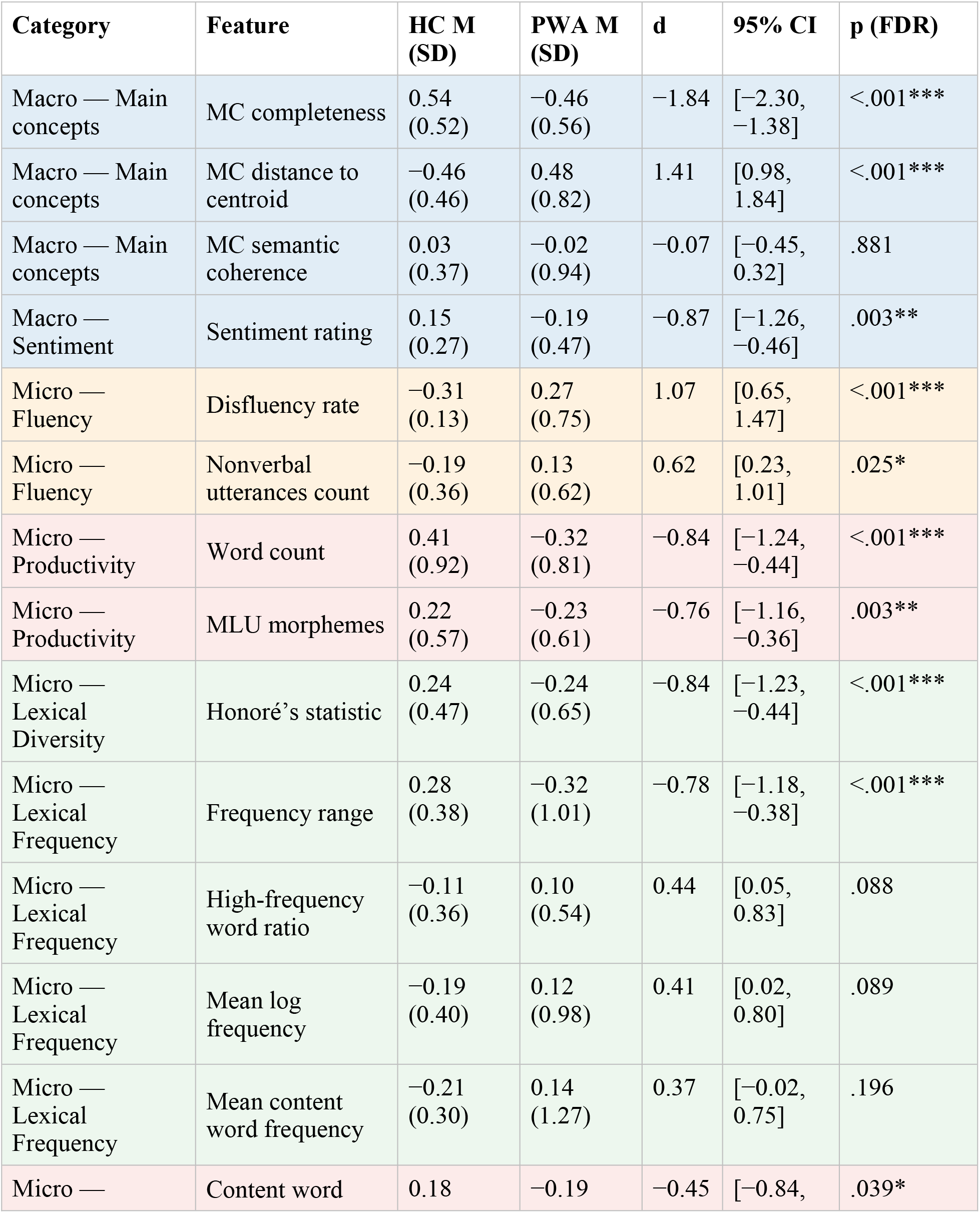
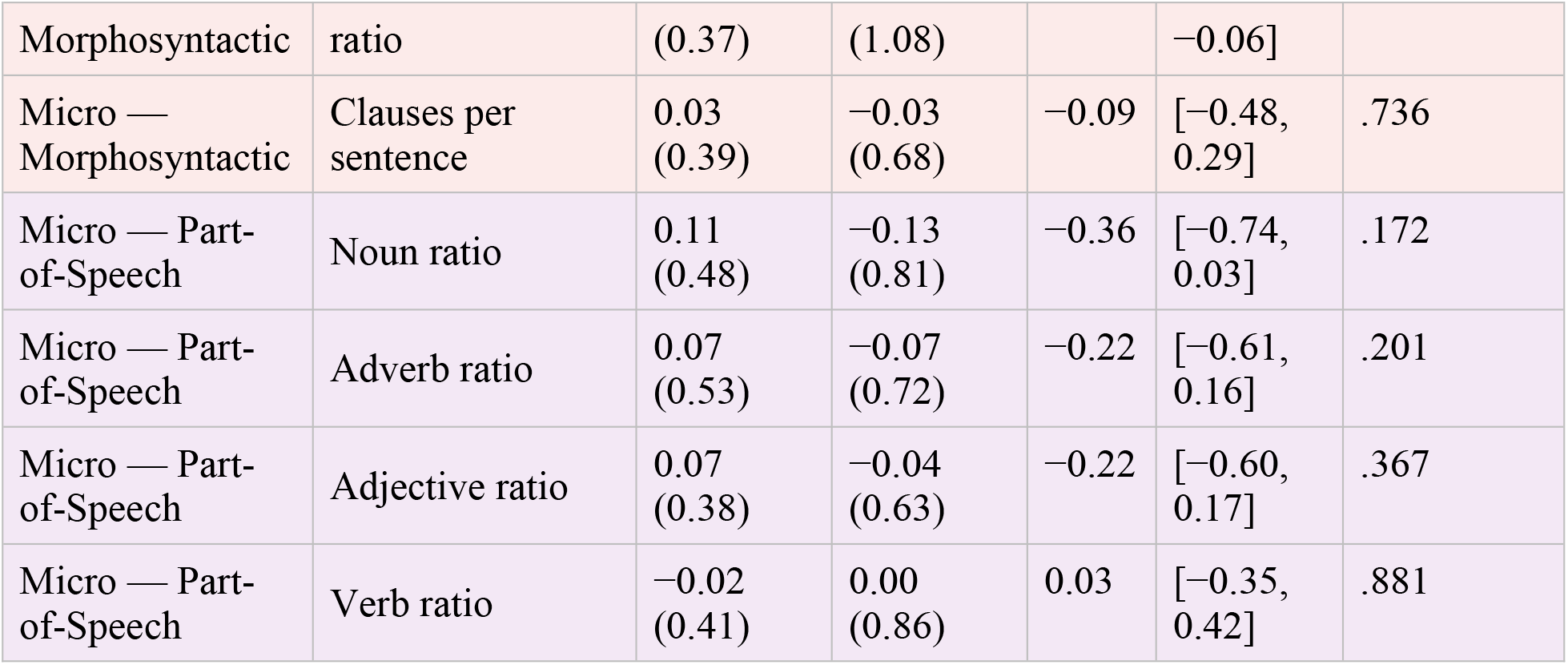
Group differences in automatic features derived from movie clip narrations. The mean (and standard deviation) of 19 macrolinguistic and microlinguistic discourse features are given in standardized z-scores for the HC and PWA groups. Group differences in these features were tested with ANCOVA controlling for age. Cohen’s d (unadjusted) with 95% confidence intervals, and ANCOVA p-values FDR-corrected for multiple comparisons, are reported; age-adjusted effect sizes and sensitivity analyses are given in Supplementary Table S8. *p < .05, **p < .01, ***p < .001. Row shading denotes feature family: blue, macrolinguistic; orange, fluency; green, lexical (diversity and frequency); pink, productivity and morphosyntactic; purple, part-of-speech.

Morphological complexity was lower in PWA (MLU morphemes: d = −0.76, p = .003). PWA produced more non-verbal utterances (d = 0.62, p = .025) and had lower content word ratios (d = −0.45, p = .039). The effect sizes for every feature and clip are given in Supplementary Table S6.

### Group Classification based on macrolinguistic and microlinguistic features

LASSO logistic regression with leave-two-out cross-validation (LTOCV; 2,754 folds) was used to classify participants based on the 19 selected discourse features derived from the movie narrations. Classification of PWA and HC was excellent (AUC = 0.948, 95% CI [0.897, 0.986]; accuracy = 0.895, 95% CI [0.838, 0.952]; sensitivity = 0.852, 95% CI [0.759, 0.944]; specificity = 0.941, 95% CI [0.882, 1.000]; Youden’s J = 0.79, 95% CI [0.68, 0.91]; Fig. 2A). Specifically, the model identified 85% of PWA and 94% of HC correctly. To estimate the contribution of each feature family, models restricted to the 15 microlinguistic or to the 4 macrolinguistic features were compared with the full model. The AUC of neither restricted model differed significantly from that of the full model (microlinguistic: AUC = 0.914, DeLong p = .070; macrolinguistic: AUC = 0.908, p = .235; Supplementary Table S9). At the cross-validation-optimal lambda, the model retained 13 of the features (Fig. 2B, D). Main concept distance to the inventory centroid had the largest coefficient of any retained feature. Ten features were selected in at least 80% of LTOCV folds and were stable contributors to classification, including two macrolinguistic features (MC completeness, MC distance to centroid) and 8 microlinguistic features (mean log frequency, nonverbal utterances count, disfluency rate, word count, adjective ratio, adverb ratio, Honore’s statistic, MLU morphemes). The other 3 features showed lower cross-fold reliability, indicating that their inclusion may not generalize. The 6 excluded features had coefficients near zero, suggesting that they did not contribute to classification. Examination of misclassified cases (Fig. 2C) indicated that of the 8 misclassified PWA, 3 had minimal or subtle language deficits (AQ ≥ 93.8, the WAB-R cut-off for the normal range), 4 had mild to moderate deficits (AQ = 73.6-92.1), and 1 had severe deficits (AQ = 26.5).

**Figure 2.**
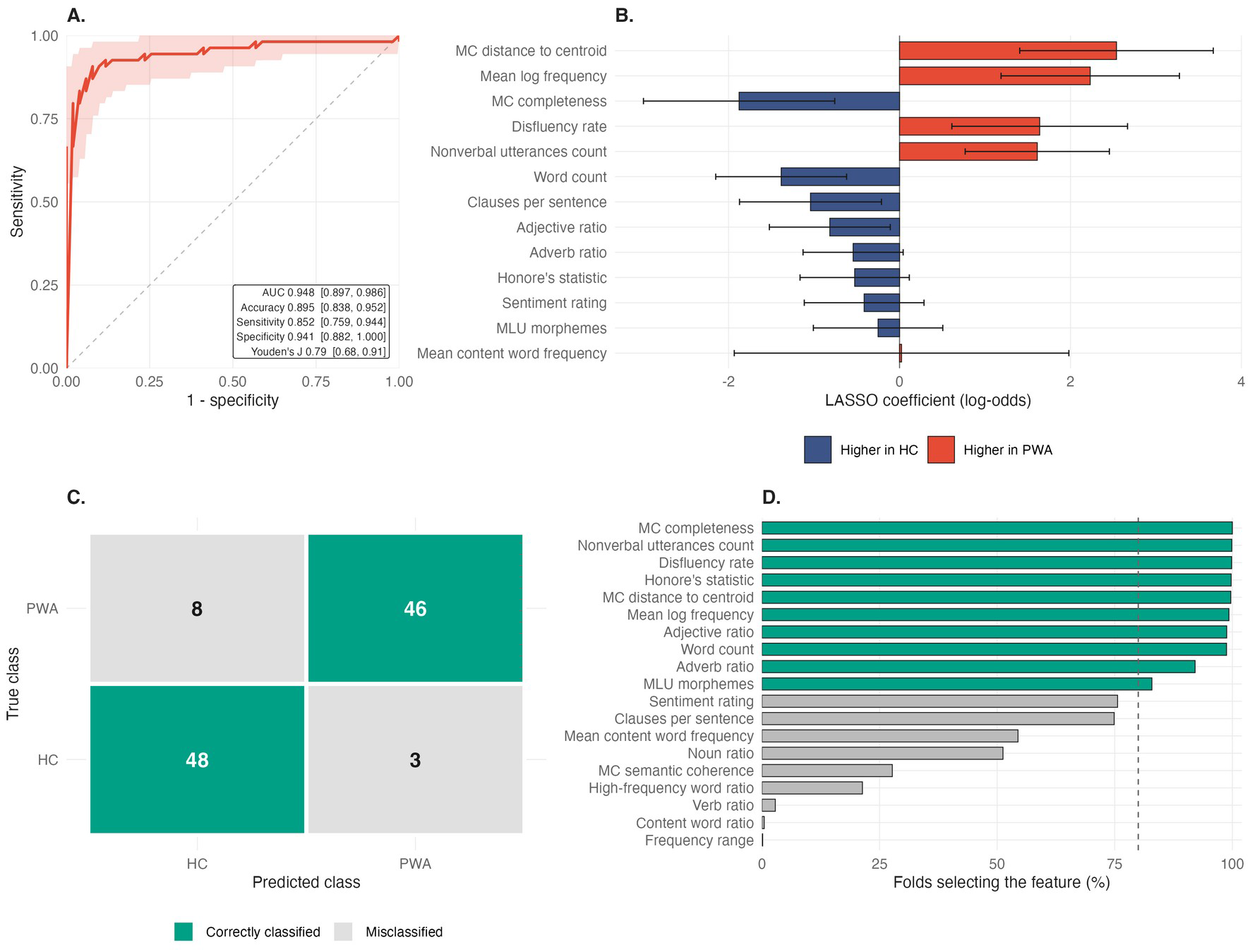
**LASSO classification performance and feature selection**. (A) Receiver operating characteristic curve from leave-two-out cross-validation. (B) Final-model coefficients ordered by absolute magnitude; a positive coefficient indicates that higher values of the feature increased the predicted probability of aphasia, and a negative coefficient that they decreased it, with the other retained features held constant (the direction of each univariate group difference is given in Table 1). Error bars are ±1 bootstrap standard error (2,000 refits at the selected penalty; Supplementary Table S11). (C) Confusion matrix: rows indicate true class and columns predicted class; fill color indicates correctly classified versus misclassified. (D) Percentage of the 2,754 folds in which each feature received a non-zero coefficient; The dashed line marks the 80% stability threshold. MC = main concept.

### Correlations of discourse scores with clinical severity

Within the PWA subsample, each discourse feature score was correlated with the Western Aphasia Battery-Revised Aphasia Quotient (AQ) (WAB-R)^10^, partialling out age and applying FDR correction for multiple comparisons. Thirteen of the 19 features were significantly associated with AQ (Table 2, Figure 3), of which 9 were among the features retained by the classification model. Of the macrolinguistic features, more severe aphasia (lower AQ) was associated with greater distance of produced concepts from the VLM-derived main concept inventory (r = −0.79, p < .001), lower main concept completeness (r = 0.66, p < .001), and more negative sentiment (r = 0.65, p < .001). The association with AQ was stronger for distance to the centroid than for completeness (Williams t = 2.14, p = .037). Of the microlinguistic features, more severe aphasia was associated with a narrower lexical frequency range (r = 0.69), lower mean log frequency (r = 0.58) and lower lexical richness (Honore’s statistic: r = 0.33), lower productivity (word count: r = 0.54; MLU morphemes: r = 0.35), and higher disfluency rate (r = −0.37). In a hierarchical regression, distance to the centroid explained AQ variance beyond age, completeness and the microlinguistic features associated with AQ (ΔR² = 0.069, 95% CI [0.001, 0.115]; F(1, 41) = 16.62, p < .001), whereas completeness explained no additional variance beyond distance (ΔR² = 0.003, p = .417; Supplementary Table S10).

**Figure 3.**
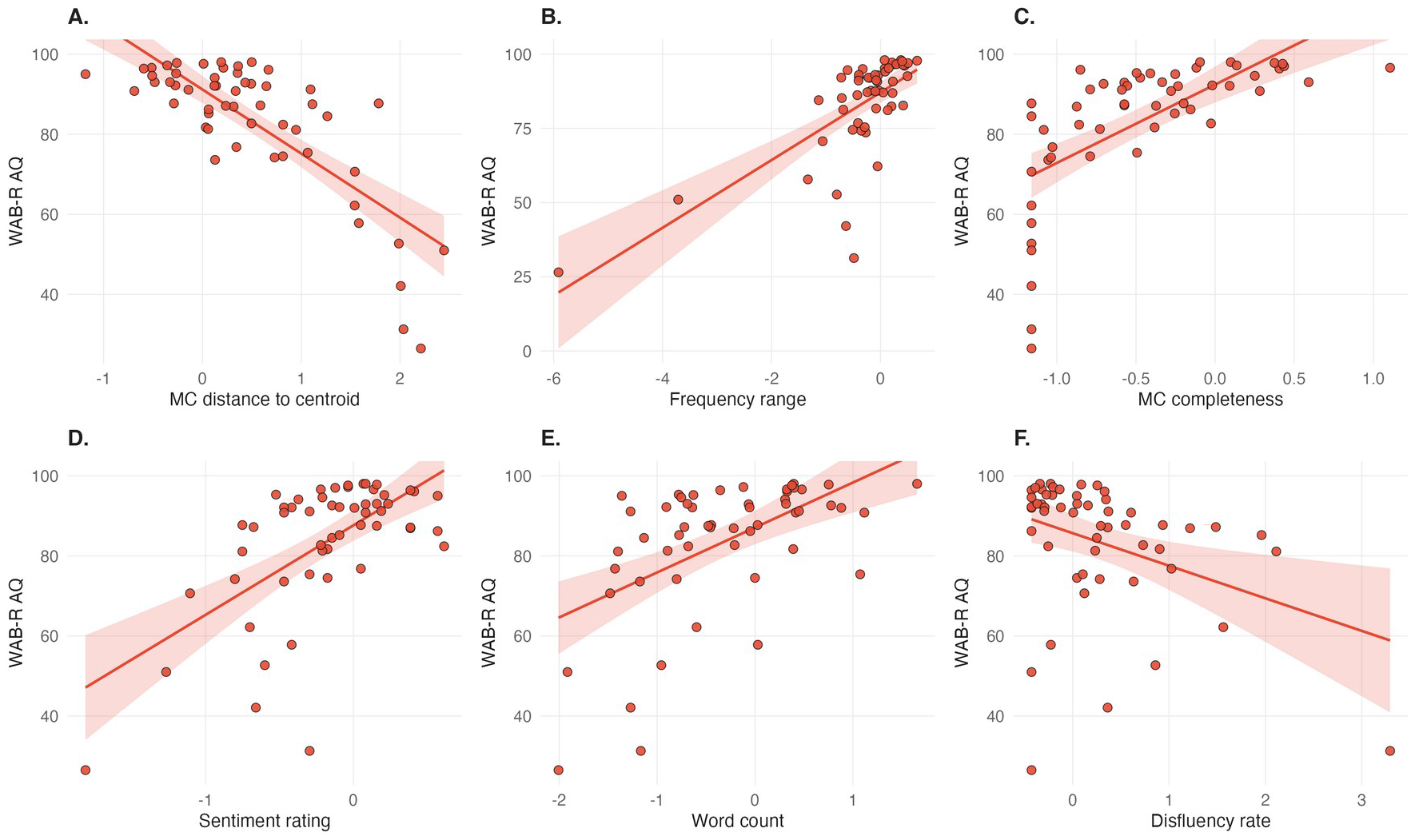
Correlations between discourse features and aphasia severity in PWA. Scatterplots showing the relationships between discourse features (z-score standardized, x-axes) and the Western Aphasia Battery-Revised Aphasia Quotient (AQ; y-axis) within the PWA subsample. The six features showing a significant association with AQ in both Pearson and Spearman partial correlations (Supplementary Table S12) are shown, ordered by absolute Pearson correlation magnitude. Higher AQ scores indicate milder aphasia. Points represent individual participants and the solid lines are linear regression fits with 95% confidence bands (shaded regions). (A) MC distance to the inventory centroid. (B) Lexical frequency range. (C) MC completeness. (D) Sentiment rating. (E) Word count. (F) Disfluency rate. MC = main concept.

**Table 2.** Correlation values between discourse features and aphasia severity. Partial correlations between all 19 discourse features and the WAB-R Aphasia Quotient in PWA, adjusted for age and FDR-corrected, ordered by absolute correlation magnitude. Shading denotes feature family: blue, macrolinguistic; orange, fluency; green, lexical (diversity and frequency); pink, productivity and morphosyntactic; purple, part-of-speech. MC = main concept; MLU = mean length of utterance.

| Feature | r (partial) | 95% CI | p (FDR) |
| --- | --- | --- | --- |
| MC distance to centroid | −0.79 | [−0.87, −0.66] | <.001 |
| Frequency range | 0.69 | [0.52, 0.81] | <.001 |
| MC completeness | 0.66 | [0.47, 0.79] | <.001 |
| Sentiment rating | 0.65 | [0.47, 0.79] | <.001 |
| Mean log frequency | 0.58 | [0.36, 0.73] | <.001 |
| Word count | 0.54 | [0.32, 0.71] | <.001 |
| Mean content word frequency | 0.42 | [0.17, 0.62] | .005 |
| MC semantic coherence | −0.40 | [−0.60, −0.14] | .008 |
| Verb ratio | 0.37 | [0.11, 0.58] | .013 |
| Disfluency rate | −0.37 | [−0.58, −0.11] | .013 |
| MLU morphemes | 0.35 | [0.08, 0.56] | .019 |
| Honoré’s statistic | 0.33 | [0.07, 0.55] | .024 |
| High-frequency word ratio | 0.32 | [0.06, 0.55] | .026 |
| Nonverbal utterances count | 0.24 | [−0.03, 0.48] | .116 |
| Adjective ratio | −0.17 | [−0.42, 0.11] | .298 |
| Content word ratio | 0.12 | [−0.15, 0.38] | .449 |
| Adverb ratio | 0.08 | [−0.20, 0.34] | .643 |
| Noun ratio | −0.06 | [−0.33, 0.21] | .699 |
| Clauses per sentence | 0.00 | [−0.27, 0.27] | .998 |

## Discussion

We evaluated whether automatically derived macrolinguistic (i.e., main concept, sentiment) and microlinguistic (i.e., speech fluency, lexical, and syntactic) features extracted from movie clip narrations can facilitate and augment the clinical assessment of discourse impairment in aphasia. Main concept inventories generated directly from each clip using a vision-language model were semantically proximal to the main concepts produced by healthy controls, supporting their utility as valid references. The macrolinguistic features showed the largest differences between PWA and HC: PWA produced fewer unique main concepts, and utterances that are semantically farther from the narrative centroid. The microlinguistic features showed more moderate group differences across domains: PWA relative to HC were less fluent and verbose, and showed lower lexical diversity and morphological complexity. LASSO classification based on the macrolinguistic and microlinguistic features achieved excellent performance, although the macrolinguistic features did not significantly improve classification beyond the microlinguistic features alone. The correlations with aphasia severity revealed that more severe aphasia was associated most strongly with greater distance of semantic concepts from the inventory and lower main concept completeness, more negative sentiment, and a narrower lexical frequency range. Thus, the automatically derived macrolinguistic features improved prediction of aphasia severity beyond the traditional microlinguistic measures alone, consistent with prior work emphasizing the clinical importance of discourse-level assessment^13,16^. The success of multimodal VLM-generated inventories as valid references supports the feasibility and scalability of automated criterion-based discourse assessment using ecologically valid stimuli, and highlights the potential of this approach for routine clinical implementation.

Our work improves computational MCA in several ways. To our knowledge, this study is the first using multimodal AI to generate main concept inventories directly from the multimodal stimulus (video, audio, subtitles) rather than just from text. Our demonstration that these inventories are acceptable approximations of healthy-control consensus concepts represents a methodological advance that will facilitate scalable adaptation to new assessment materials, bypassing the need for iterative expert derivation of an inventory for each stimulus. The VLM enabled retrieval of concepts grounded strictly in audiovisual content, e.g., for the dialogue-free animated clip. These concepts were produced by HC at rates comparable to the subtitle-grounded concepts, further emphasizing the relevance of the multimodal MCA approach. Fabricated content is a documented failure mode of vision-language models^22,23^; here, each concept had to cite a subtitle quote or a frame timestamp from the clip, and concepts that fewer than 20% of HC produced were removed. In addition, the stimuli used here for eliciting the narrations were relatively long (2 to 5 minutes) movie clips with multiple connected scenes, lending higher ecological validity to the narrative assessment than the brief BATS videos used in a prior MCA study^18,19^, and the single static pictures dominating standardized aphasia batteries^11^.

Among the main concept features, completeness showed the larger group difference, whereas semantic distance to the centroid was more strongly associated with aphasia severity and explained severity variance beyond completeness. The correlation strength of the main concept semantic distance feature with aphasia severity also exceeded that reported in prior manual MCA studies using a categorical concept completeness measure^4,13,16^. Thus, the embedding-based distance-to-centroid feature may preserve graded severity information that is lost in the traditional ‘concept present/absent’ scoring. This finding reinforces the value of the embedding semantic measure for accurately capturing narrative processing performance, and highlights its potential for identifying new and concrete intervention targets.

Narrative sentiment also emerged as an informative macrolinguistic feature that differentiated between the groups and was correlated with aphasia severity. The more negative narrative sentiment observed in the aphasia versus control group may reflect difficulties in narrative comprehension and production, and/or difficulties in emotion perception and expression. The finding demonstrates the feasibility of computing narrative sentiment at scale using a large language model^24^. The finding also highlights the potential value of the sentiment measure for characterizing aspects of narrative processing which may affect functional communication in aphasia and are seldom considered.

Movie narration approximates the demands of everyday storytelling and social interaction, dynamically and simultaneously engaging linguistic, multimodal sensory, attentional, working memory, and motor systems essential for functional communication^11,25^. The movie narration probe may therefore predict real-world communicative success more accurately than decontextualized tasks (e.g., Cookie Theft, confrontation naming, sentence repetition) which lack the temporal structure and social-pragmatic demands of connected discourse. Contemporary treatment frameworks for aphasia prioritize functional communicative success over impairment-level metrics^11^, as stated in the WHO International Classification of Functioning, Disability and Health^26^ and operationalized in frameworks such as the Life Participation Approach to Aphasia^27^ and A-FROM^25^. The clinical utility of functional features spans diagnosis, severity assessment, and treatment monitoring. Analysis of the LASSO classification model relative to aphasia severity indicates that the discourse features capture language impairment even at AQ ≥ 93.8 (12 of the 15 PWA in that range were correctly classified), where the WAB-R test no longer distinguishes individuals with aphasia from neurotypical controls^10^, suggesting sensitivity of the movie narration probe at the mild end of the severity spectrum where standardized batteries lose resolution.

## Conclusions and Limitations

Main concept inventories generated directly from multimodal movie input using a VLM approximated the main concepts healthy speakers produced, making them adequate to serve as reference inventories for computerized main concept analysis. This methodological advance alleviates a norming bottleneck that has long constrained main concept analysis to a small set of (often outdated) pictures and stories. The new methodology opens a path to libraries of discourse assessment stimuli that are more realistic, diverse, and adaptable to varying cultural and clinical needs. Furthermore, the study demonstrated that automatic main concept features derived from the movie clip narrations, and in particular the semantic distance from the inventory centroid, contained signal relevant to accurate aphasia classification and prediction of severity, beyond the traditional categorical main concept, and microlinguistic, measures alone. This finding is consistent with longstanding clinical evidence that aphasia disrupts narrative informativeness and coherence independently of fluency and syntax^13,16^, lending support to the validity and utility of the automatic discourse features, and the current emphasis on rehabilitation frameworks that prioritize functional communication^25,27^. Overall, the findings support the feasibility and scalability of automated criterion-based discourse assessment using ecologically valid stimuli, and highlights the potential of this approach for routine clinical implementation.

Several limitations warrant consideration. First, while the VLM-generated inventories corresponded to HC-produced main concepts, they have not been externally validated against the consensus of aphasiologists who are experts in manual MCA inventory development^4,16^. A direct comparison with expert-generated main concepts for these movie clips would further strengthen confidence in the clinical meaningfulness of the inventories. Second, the findings are specific to Gemini 2.5 Pro and may not generalize to other VLMs. Kurland et al.^18^ found that both proprietary (GPT-4) and open-source (Llama-3-70B) models achieved strong agreement with human main concept scoring, suggesting some stability for the scoring task. Whether similar cross-model consistency is maintained for multimodal inventory generation requires empirical verification. Third, the aphasia sample was composed mostly of anomia, precluding an analysis of aphasia subtypes. Dalton and Richardson^13^ reported varying effect sizes for MCA measures across aphasia subtypes and discourse tasks. Future research with larger samples, stratified by aphasia type, can determine whether automated MCA features derived from movie narrations show selective sensitivity to particular language impairment patterns and clinical profiles.

Finally, correspondence between the inventories and HC production varied about twofold across clips (mean 30% to 62% of inventory concepts produced by HC; Supplementary Table S2), suggesting that the adequacy of a VLM-generated inventory likely depends on the stimulus characteristics.

## Methods

### Ethical Considerations

The study was approved by the Boston University Charles River Campus Institutional Review Board (Protocol #3309E) and the Mass General Brigham Institutional Review Board (Protocol # 2023P002772). All participants provided written informed consent prior to enrollment and were compensated for their participation. No participant data were shared with commercial entities.

PWA completed neuropsychological assessment in a remote session (1 hour) that included the WAB-R^10^, based on which the aphasia quotient (AQ) is computed. The movie-viewing paradigm (2 hours) was completed in a second session conducted remotely (n = 36 PWA) or in person (n = 18 PWA) to accommodate different participant needs. HC participants completed a single in-person session (2 hours) beginning with Montreal Cognitive Assessment (MoCA)^28^ screening, followed by the movie-viewing paradigm. Participants with MoCA scores falling in the mild cognitive impairment range (<25, n=17) were excluded from this study as their movie narrations may be less representative of normal discourse. Five to eight clips were presented in randomized order, depending on the time available in the session. Before presenting each clip, the experimenter provided a one-sentence neutral description (e.g., “This clip features a conversation between two men in a restaurant”). After viewing each clip, participants were instructed to answer a few content questions, and then provide a free narration using the prompt: “Tell me in your own words what happened in the clip.” (limited to 60 seconds). The narrations were audio-recorded for subsequent transcription and analysis. Other procedural details are given in ref. ^29^.

### Participants

The characteristics of the sample analyzed here, are summarized in Table 3.

**Table 3.** Sample characteristics. HC = healthy controls; PWA = persons with aphasia; M = mean; SD = standard deviation; n = number; WAB-R = Western Aphasia Battery–Revised; AQ = Aphasia Quotient; MoCA = Montreal Cognitive Assessment.

| Characteristic | HC (n = 51) | PWA (n = 54) |
| --- | --- | --- |
| Age, M ± SD (range) | 48.9 ± 16.0 (22–69) | 59.5 ± 10.6 (38–80) |
| Gender, n (%) | Female: 29 (56.9%)<br>Male: 22 (43.1%) | Female: 23 (42.6%)<br>Male: 31 (57.4%) |
| Race, n (%) | Asian: 7 (13.7%) | Asian: 5 (9.3%) |
|  | Afr. American: 5 (9.8%)<br>White: 39 (76.5%)<br>Unknown: 0 (0%) | Afr. American: 6 (11.1%)<br>White: 42 (77.8%)<br>Unknown: 1 (1.9%) |
| Ethnicity, n (%) | Hispanic/Latino: 2 (3.9%)<br>Not Hispanic/Latino: 49 (96.1%)<br>Unknown: 0 (0%) | Hispanic/Latino: 2 (3.7%)<br>Not Hispanic/Latino: 51 (94.4%)<br>Unknown: 1 (1.9%) |
| MoCA, M ± SD (range) | 28.2 ± 1.3 (26–30) | — |
| Months post aphasia onset, M ± SD (range) | — | 90.5 ± 89.2 (6–551) |
| WAB-R AQ, M ± SD (range) | — | 83.5 ± 16.7 (26.5–98) |
| Aphasia type, n (%) | — | Anomic: 38 (70.4%)<br>Broca's: 5 (9.3%)<br>Conduction: 3 (5.6%)<br>Not Aphasic (by WAB): 6 (11.1%)<br>Transcortical Sensory: 2 (3.7%) |

### Stimuli

Seven short clips (140–348 seconds) from well-known live action movies, featuring dialogue-driven plots with clear story lines, were selected from the Dynamic Affective Movie Clip Database for Subjectivity Analysis (DynAMoS), a corpus of emotionally evocative film excerpts^30,31^. An eighth clip consisting of a wordless animated short (348 seconds) was included to examine narrative processing independent of language comprehension. The clips were extracted from high-definition source material, and presented at 1920×1080 resolution, with audio levels normalized between clips, and without subtitles to avoid confounding effects of reading ability (additional details in refs. ^29,30^).

### Transcription

The audio recordings were transcribed using CrisperWhisper 1.0^32^, an open-source automatic speech recognition model optimized for verbatim transcription with standardized annotation of disfluencies (non-verbal utterances, repetitions, false starts), and with accurate word-level onset and offset timestamps. The model was accessed via the Hugging Face repository (nyrahealth/CrisperWhisper, the original v1 release, fine-tuned from Whisper large-v3) and run locally to ensure data privacy and HIPAA compliance. Each transcript was manually reviewed, and if needed, corrected by the first author (M.J.M.), a licensed speech-language pathologist, to ensure accuracy of lexical content, appropriate segmentation of utterances, and correct identification of disfluencies. The fourth author (K.G.), a trained research assistant, independently reviewed 25% of the transcripts from each group to spotcheck transcription accuracy.

### Feature Extraction

#### Microlinguistic Features

Microlinguistic features were extracted using the *dpfluency* module of the *dptools* Github repository, a computational linguistics toolkit for speech and discourse analysis. Additional libraries used included Stanza^33^ for part-of-speech tagging and utterance segmentation, and Sentence-Transformers^34^ for semantic embeddings. The extracted features spanned six domains: productivity (word count), morphosyntax (mean length of utterance in morphemes, clauses per sentence, content word ratio), lexical diversity (Honoré’s statistic), lexical frequency (mean log frequency, frequency range, high-frequency word ratio, mean content word frequency; derived from COCA^35^), fluency (disfluency rate, nonverbal utterances count), and part-of-speech distribution (noun, verb, adjective, and adverb ratios). The features were computed as described in Liebenthal et al.^21^ and averaged across utterances within each movie-clip narration transcript. Detailed feature definitions are provided in Supplementary Table S4.

#### Macrolinguistic Features: Main concepts

Main concept inventories were generated for each movie clip with Gemini 2.5 Pro^36^, a vision-language model, from the clip’s video frames, audio, and subtitles. The pipeline is shown end to end in Supplementary Figure S1. Each clip was preprocessed to extract representative frames using a hybrid of scene-change detection (threshold 0.30) and uniform temporal sampling at approximately one frame every five seconds, capped at 40 frames per clip. Mono audio was sampled at 16 kHz along with a time-aligned transcript of the dialogue (subtitles). These inputs and the clip title were supplied with a structured prompt built on MCA conventions^16^ and narrative role principles^37^, under four constraints: (1) no world knowledge about the source films beyond what the clip presents, other than obvious inferences about the scene, (2) character names allowed only if spoken in the clip, (3) each proposed concept required supporting evidence from the clip itself, in the form of a subtitle quote and/or a video frame timestamp, so that every concept could be traced to the stimulus, and (4) coverage of events was required across the beginning, middle, and end of the clip, including its outcome and events shown but not spoken.

Between 5 and 15 concepts were returned per clip, each with a one-sentence description and a narrative role label.

Because VLM decoding is stochastic^38^, five sets of concepts were generated independently per clip and treated as a panel from which a single inventory was assembled. Concept labels from all five runs (271 across the eight clips) were embedded with all-mpnet-base-v2^34^. Concepts from different runs were treated as the same concept when the cosine similarity of their labels exceeded 0.63, the 95th percentile of similarity between concepts that a single run had listed as distinct, and the cluster medoid was retained as the concept label.

Concepts represented in at least three of the five runs per clip were retained, yielding 54 concepts across clips. As a final step, concepts produced by fewer than 20% of the HC sample were excluded because they were considered less relevant to discourse assessment. For this step, a control was counted as producing a concept when it was nearest the inventory concept at cosine similarity above 0.50. Fifteen concepts were excluded based on low HC production, yielding a total of 39 concepts across eight clips in the final inventories (Supplementary Table S1). Because the control narrations that set this criterion are also the control group in the analyses, all participants were also scored against the unfiltered 54-concept inventory as a sensitivity analysis (Supplementary Table S13).

The participants’ movie narrations were analyzed with respect to the main concept inventories, using an automated main concept analysis pipeline originally developed for the Cinderella Story^20^ and adopted here. For each clip, a centroid embedding representing the semantic core or “gist” of the narrative was computed based on the main concept inventory, using the all-mpnet-base-v2 sentence transformer^34^. The participant discourse samples were segmented into utterances (consisting of subject-verb units that may span multiple clauses) using Stanza part-of-speech tagging^33^ and pattern-based subject-verb detection. The semantic embeddings derived with all-mpnet-base-v2^34^ and normalized to unit length, and the other features, were computed per utterance and then aggregated across the narrative. Three main concept features were computed: (1) *Main concept completeness*: Each utterance was assigned to a single nearest inventory concept based on cosine similarity, and a concept was counted as produced when similarity exceeded 0.50. Completeness was computed as the proportion of a clip’s inventory concepts that was produced (see Supplementary Table S7 for results using thresholds from 0.40 to 0.70). (2) *Main concept distance to centroid*: Semantic distance was computed as the mean cosine distance (1 minus cosine similarity) from each of the participant’s utterances to the inventory centroid. Lower values indicate greater similarity to the narrative’s semantic “gist”. For visualization (Figure 1B), each participant’s mean utterance embedding for a clip, together with that clip’s inventory concepts and the inventory centroid, was projected to two dimensions by metric multidimensional scaling of their pairwise cosine distances fitted separately per clip. (3) *Main concept semantic coherence*: Coherence was computed as the mean cosine similarity between consecutive utterance embeddings in a narration, indexing the smoothness of semantic transitions.

### Macrolinguistic Features: Sentiment

Narrative sentiment was included as an index of narrative processing, since verbally conveying a story’s emotional tone and salience depends on narrative comprehension and expression abilities. The emotional valence of a participant’s retelling provides a content-level narrative processing measure, albeit coarse as it is summarized across the entire narration of each clip. The emotional valence of each discourse sample was estimated using the Gemma 3 (12B) large language model^39^, following prior validation work that demonstrated good agreement of this method with human ratings^24^. The model was run locally using LMStudio software (version 0.3.5) to maintain data security. Sentiment ratings on an ordinal scale from 1 (very negative) to 7 (very positive) were obtained using zero-shot prompting, following the procedure and prompt reported by Girard et al.^24^. Sentiment scores were z-standardized across all participants, as were the other features.

### Data Preprocessing

Features were extracted from narrative transcripts of all participant-clip combinations, resulting in 688 observations (326 from HC, 362 from PWA), representing 6.6 ± 1.4 clips completed per participant on average (range: 2-8 clips). Preprocessing was designed to reduce dimensionality while preserving clinically meaningful variance. Starting with 36 features, we first removed 3 redundant derivative summary statistics (maximum, minimum and standard deviation of syllable count per utterance). We then computed pairwise Pearson correlations among the remaining 33 features and removed one member of every pair correlating at |r| > 0.60, resulting in 19 retained features. This step was applied to reduce collinearity while retaining features from every domain. The retained features were z-score standardized using the scale() function in R. The final feature set comprised 15 microlinguistic features and 4 macrolinguistic features, as listed and defined in Supplementary Table S4.

### Statistical Analyses

The analyses were conducted in R version 4.4.2^40^, using glmnet 4.1.8 for the penalized regression, pROC 1.18.5 for the receiver operating characteristic analysis, ppcor 1.1 for partial correlations, and effectsize 1.0.0 for Cohen’s d, with significance set at α = .05. Embedding and main concept scoring ran in Python 3.11.13 with sentence-transformers 5.1.0, PyTorch 2.5.1, and NumPy 2.2.6. For analyses with multiple comparisons, we applied False Discovery Rate (FDR) correction^41^. For group comparisons, partial correlations and classification analyses, the features were aggregated to participant-level means across the clips each participant completed (PWA 6.7 ± 1.4 clips, HC 6.4 ± 1.4). Session modality (in-person vs. remote), adjusted for age and AQ, did not significantly predict word count (β = −0.22, p = .29) or MLU (β = −0.33, p = .06) (see Supplementary Table S5), and was not included as a covariate in the primary models.

#### Group comparisons

Group comparisons on discourse features used ANCOVA with age as a covariate, given the significant age difference between the groups. We compared effect size magnitudes between macrolinguistic and microlinguistic measures using bootstrap resampling (2,000 iterations, stratified by group) to construct 95% confidence intervals for the difference in mean |d|. Statistical significance was assessed via an exact permutation test (all 3,876 possible assignments of the 19 features to the macrolinguistic and microlinguistic sets).

#### Classification

We used LASSO (Least Absolute Shrinkage and Selection Operator)^42^ logistic regression to estimate group classification (PWA, HC), with leave-two-out cross-validation (LTOCV). In each of 2,754 folds, one PWA and one HC were held out, with the model trained on the remaining 103 participants and λ selected via nested 5-fold cross-validation on the training set.

Predicted probabilities were averaged across all folds in which each participant appeared. To estimate the contribution of the macrolinguistic and microlinguistic features, models were fitted to the full feature set, to the 15 microlinguistic features alone, and to the 4 macrolinguistic features alone, and the nested models were compared with DeLong’s test for correlated ROC curves^43^; two further models used age alone and the full feature set with age as an unpenalized covariate. Classification performance was evaluated using AUC, accuracy, sensitivity, specificity, and Youden’s J statistic (sensitivity + specificity − 1). Coefficients were taken from the final model refitted on all data, with lambda selected by averaging the cross-validation curve over 100 repeated 5-fold splits. Feature stability was assessed by calculating selection frequency across LTOCV folds. Features with non-zero coefficients were considered to contribute to classification. Coefficient uncertainty was estimated by refitting the model on 2,000 bootstrap resamples at the selected penalty (Supplementary Table S11).

#### Associations with aphasia severity

Within the PWA subsample, feature–AQ associations were estimated using partial correlations adjusting for age. The correlations of the two main concept features with AQ were compared with the Williams test for dependent correlations^44^. To test whether semantic distance predicts severity beyond categorical scoring, AQ was regressed hierarchically on age, the nine microlinguistic features significantly associated with AQ (Table 2), main concept completeness and main concept distance to the centroid, entered in that order and with the two main concept features also entered in the reverse order; the 95% confidence interval for ΔR² was obtained from 2,000 bootstrap resamples of participants.

## Data availability

The datasets generated and analyzed during this study are not publicly available due to privacy restrictions and IRB requirements protecting participants with neurological conditions.

Deidentified linguistic features and aggregate statistics presented in this manuscript are available from the corresponding author upon reasonable request and execution of an appropriate data use agreement. The automatically generated Main Concept Inventories for all 8 movie clips are provided in the Supplementary Materials (Supplementary Table S1).

## Code availability

Source code for the automated main concept analysis pipeline is available at https://github.com/mjmarte/mca-movie-discourse. The repository includes preprocessing scripts, feature extraction modules, the inventory construction and scoring pipeline, and the prompts used for inventory generation and sentiment rating.

## Authors’ Contributions

MJM: Conceptualization, Data curation, Formal analysis, Investigation, Methodology, Project administration, Software, Validation, Visualization, Writing—original draft, Writing—review and editing

SL: Methodology, review and editing

SW: Methodology, review and editing

MV: Methodology, review and editing

KG: Data curation, review and editing

JMG: Resources, methodology, review and editing

RB: Data curation, Investigation, review and editing

XW: Data curation, review and editing

JTB: Resources, methodology, review and editing

YT: Funding acquisition, review and editing

SK: Conceptualization, funding acquisition, methodology, resources, supervision, writing, review and editing

EL: Conceptualization, funding acquisition, methodology, resources, supervision, writing, review and editing

## Data Availability

The datasets generated and analyzed during this study are not publicly available due to privacy restrictions and IRB requirements protecting participants with neurological conditions. Deidentified linguistic features and aggregate statistics presented in this manuscript are available from the corresponding author upon reasonable request and execution of an appropriate data use agreement. The automatically generated main concept inventories for all eight movie clips are provided in the Supplementary Materials (Supplementary Table S1). Analysis code and prompts are available at https://github.com/mjmarte/mca-movie-discourse.

## Supplementary Information

Abbreviations: AQ, Aphasia Quotient; CI, confidence interval; HC, healthy controls; VLM, vision-language model; MC, main concept; PWA, persons with aphasia.

**Table S1.**
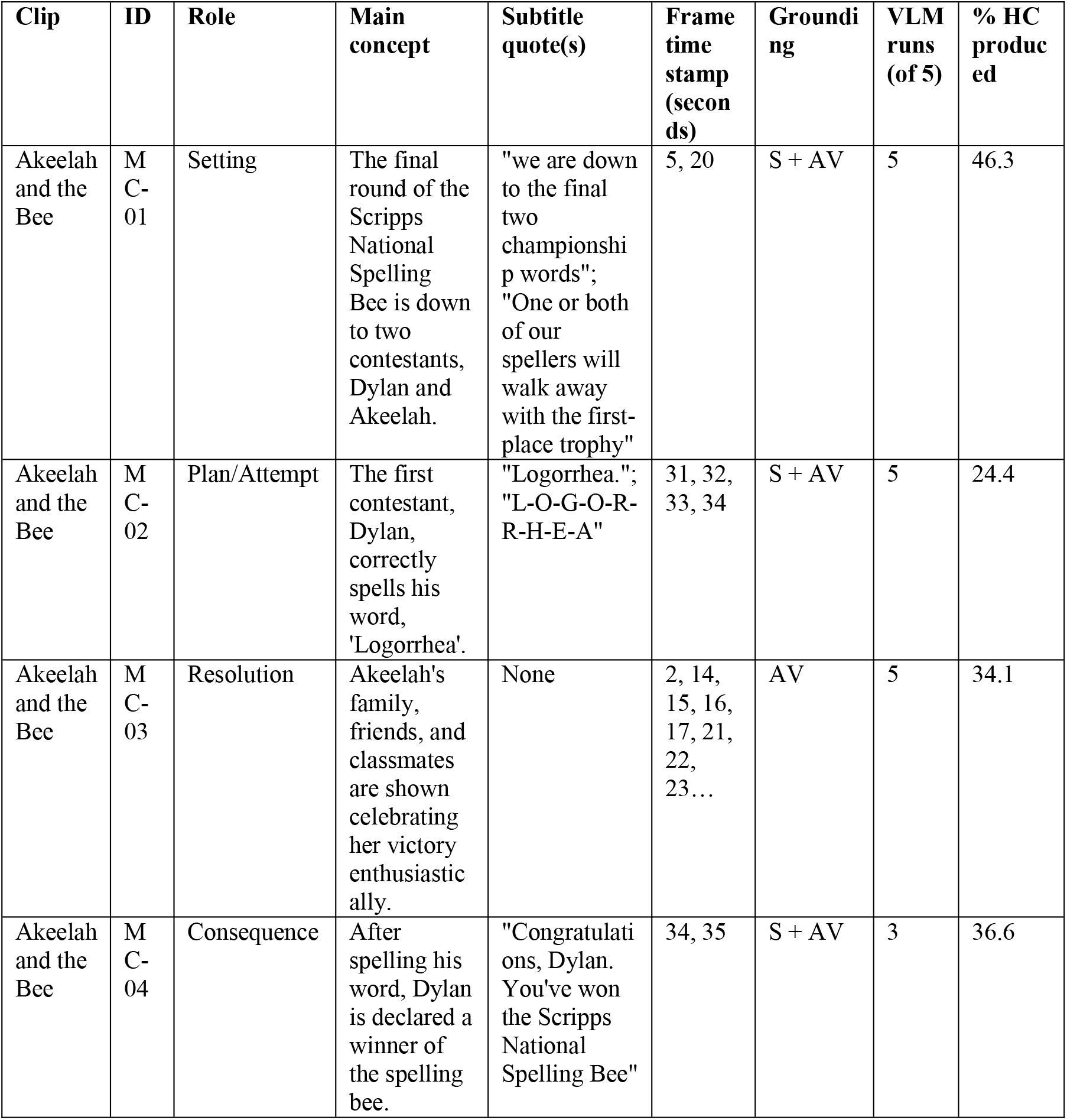
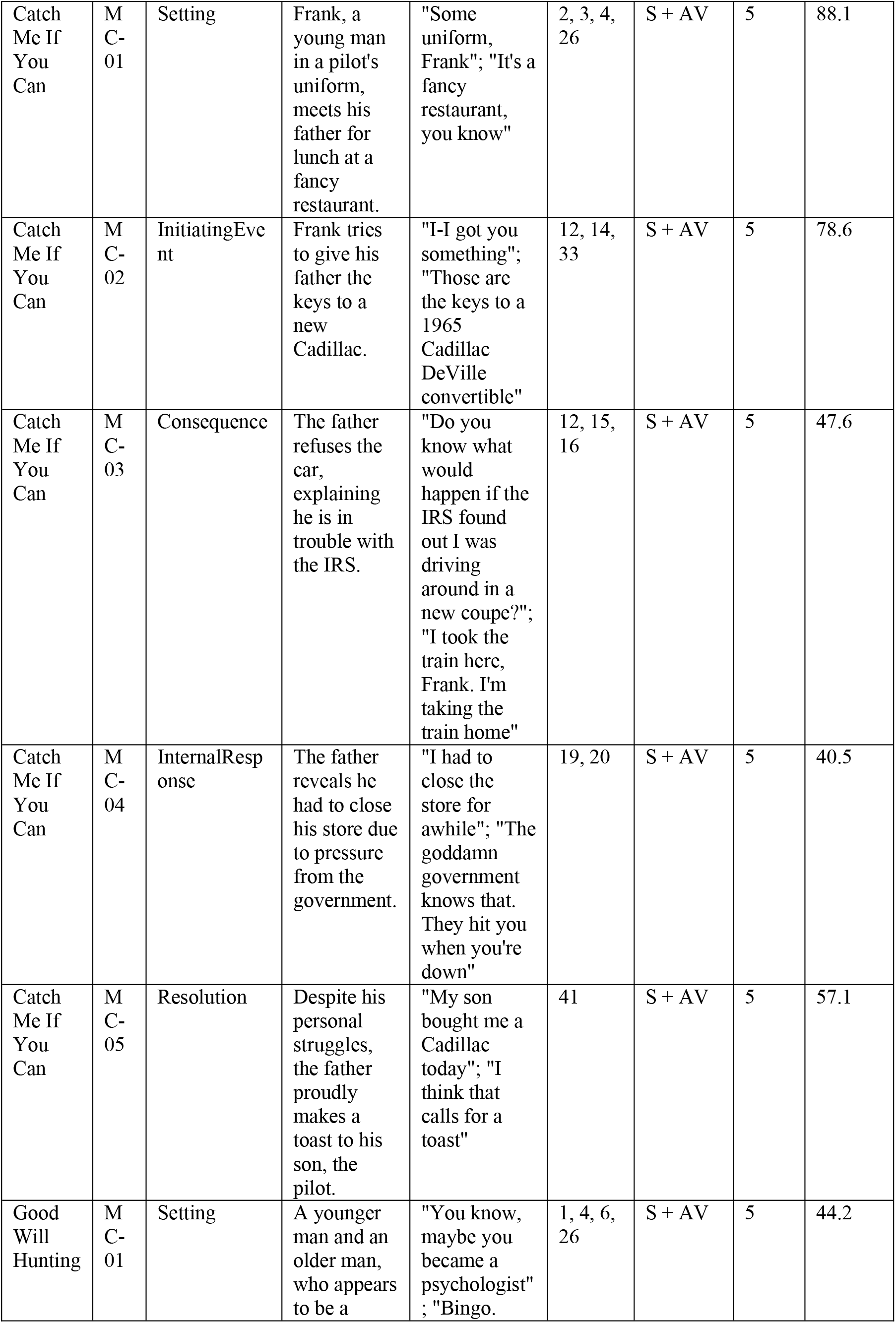
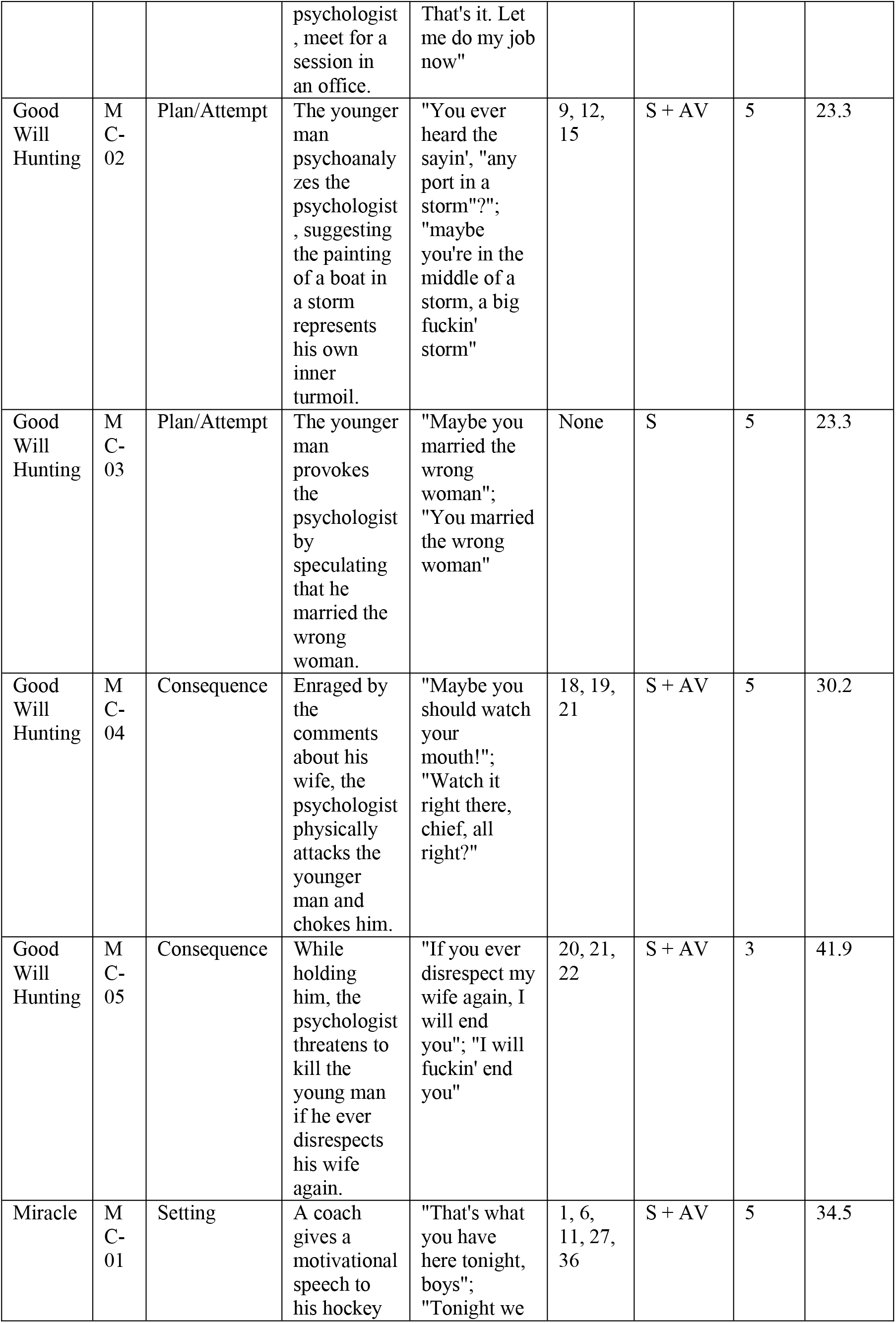
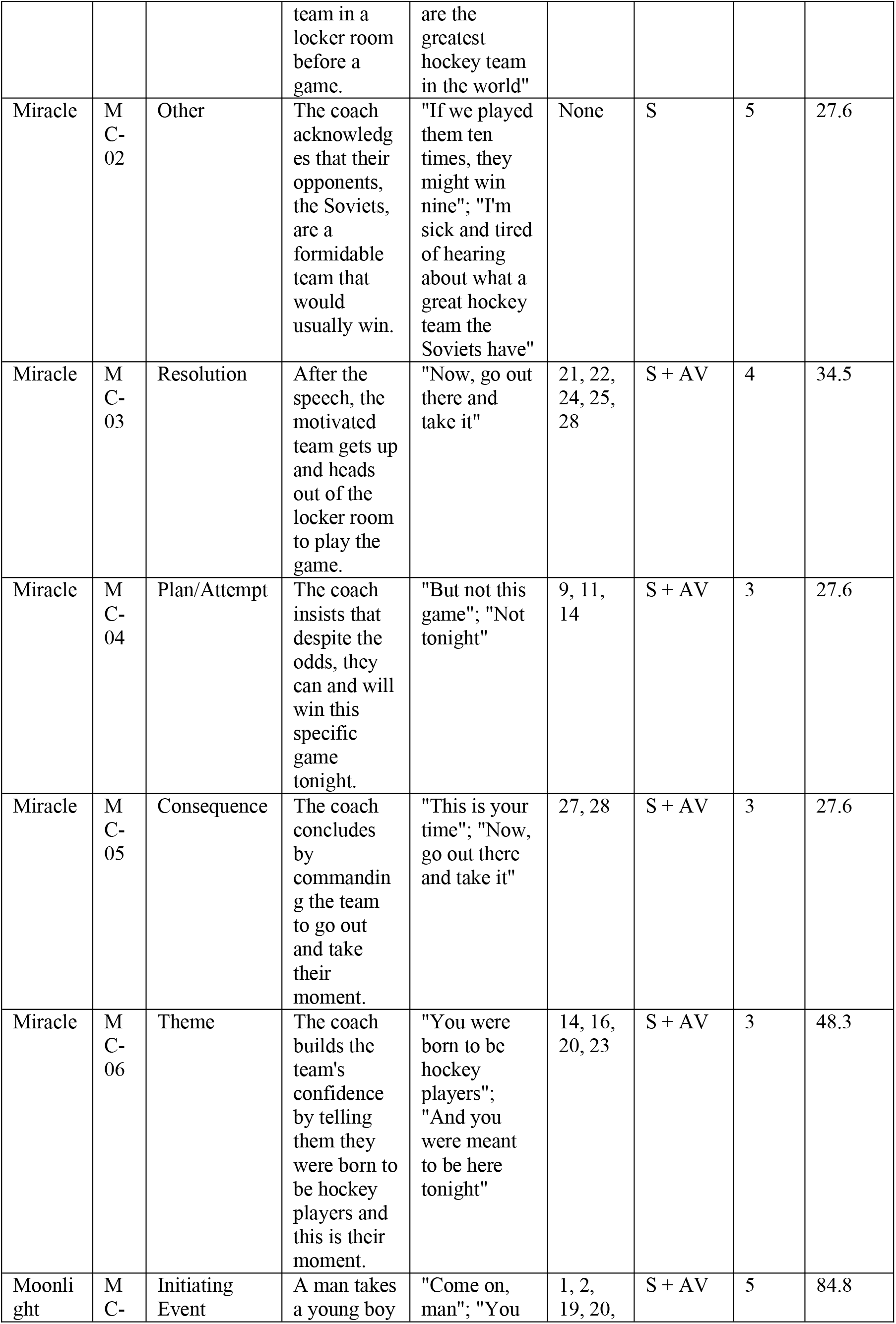
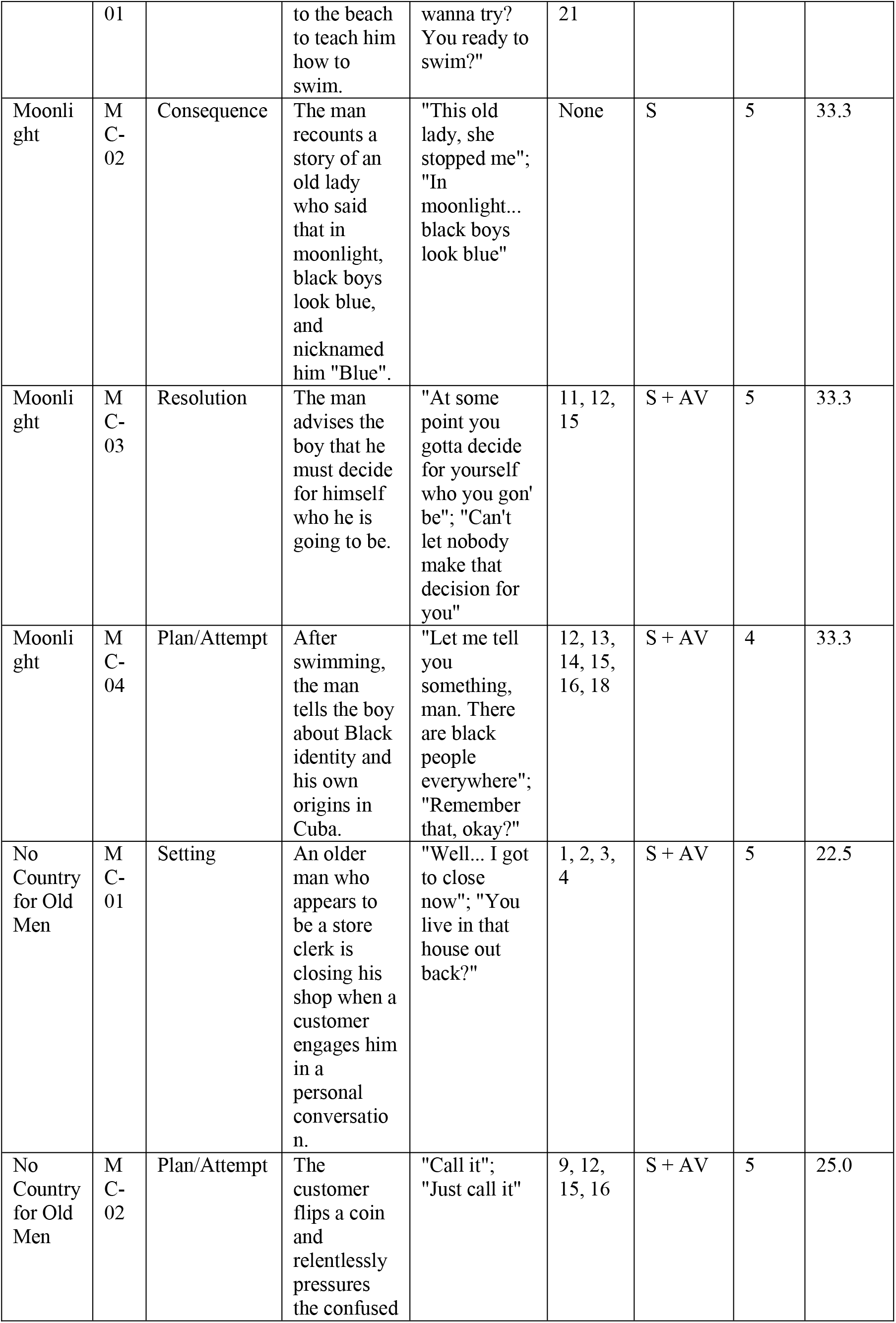
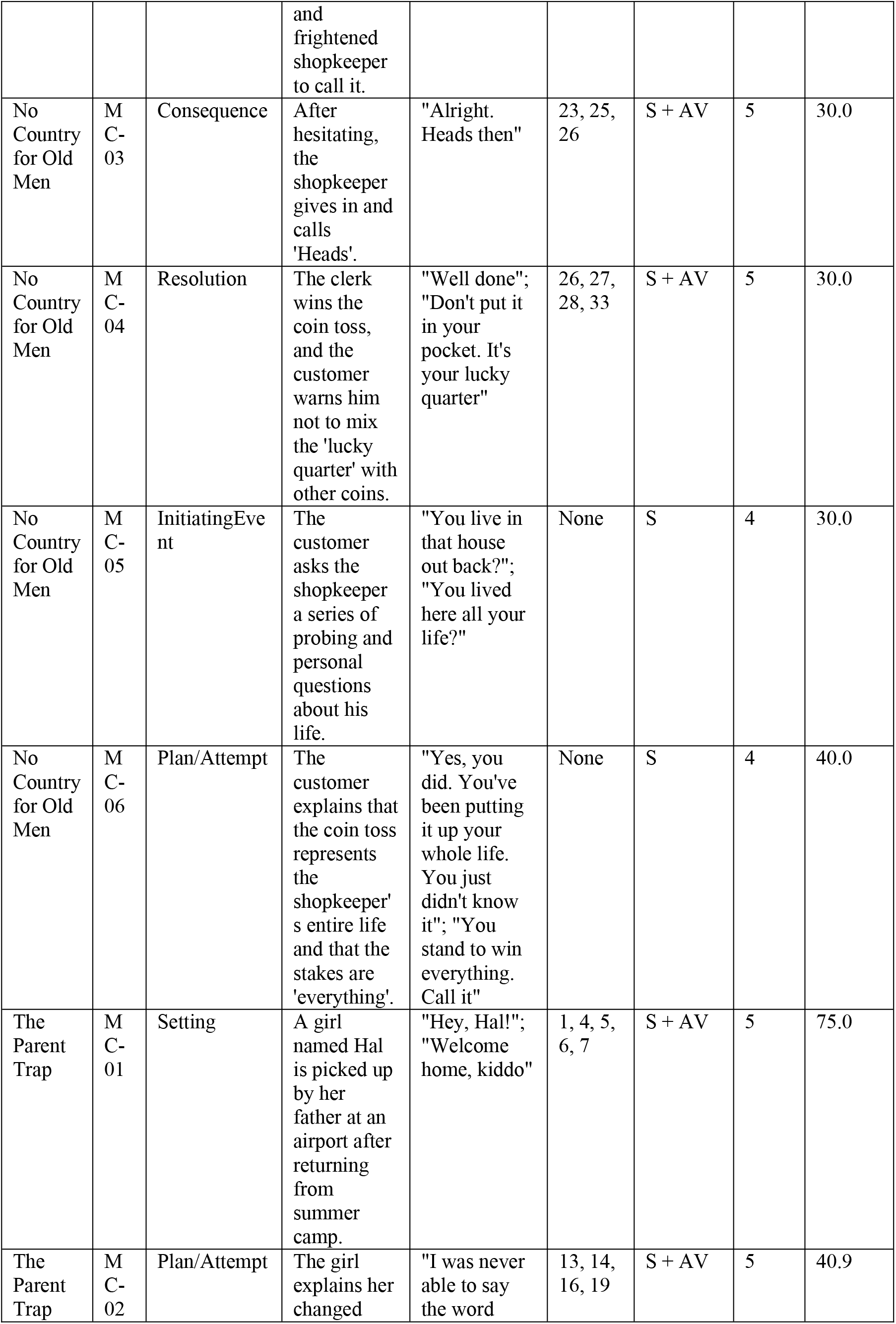
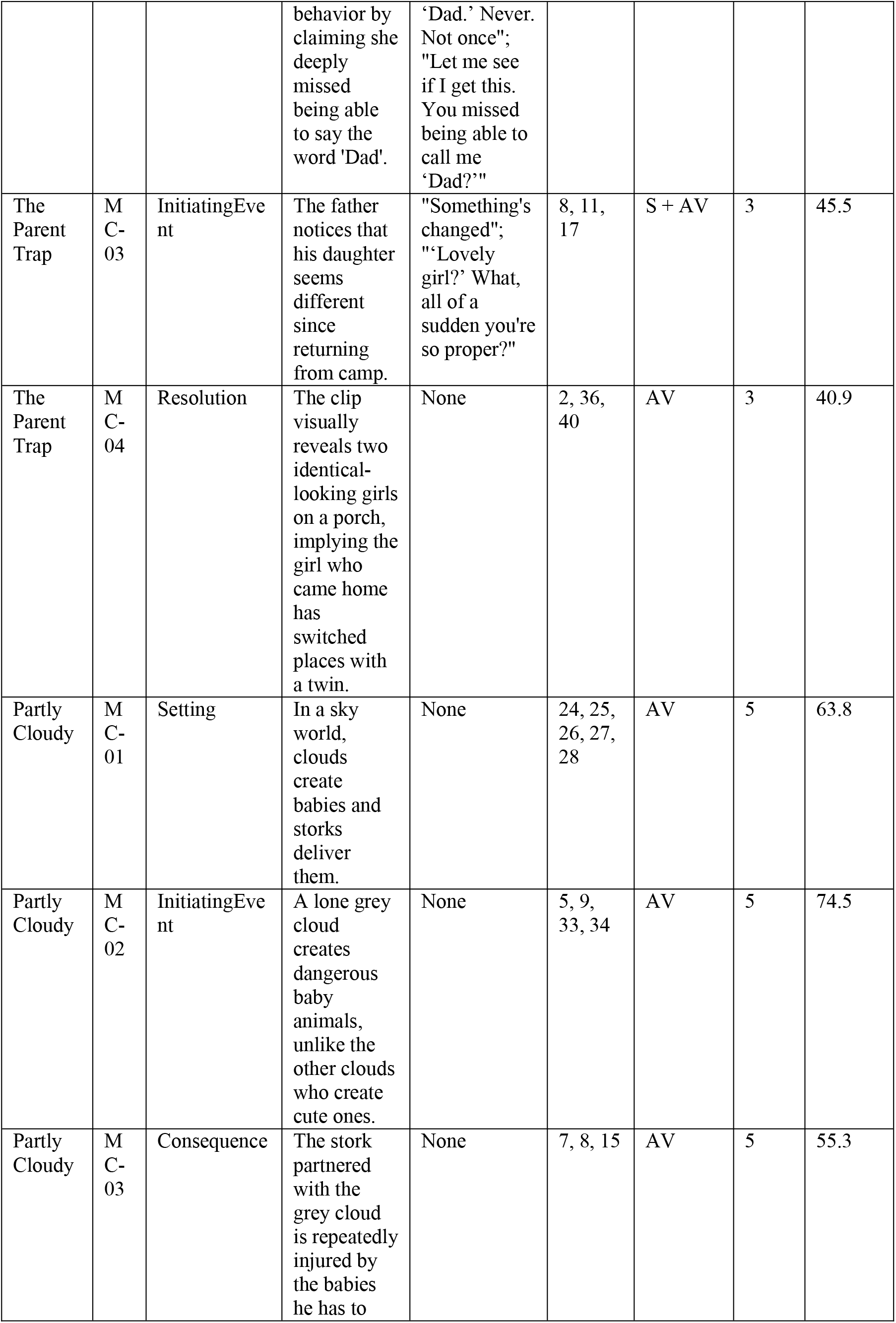
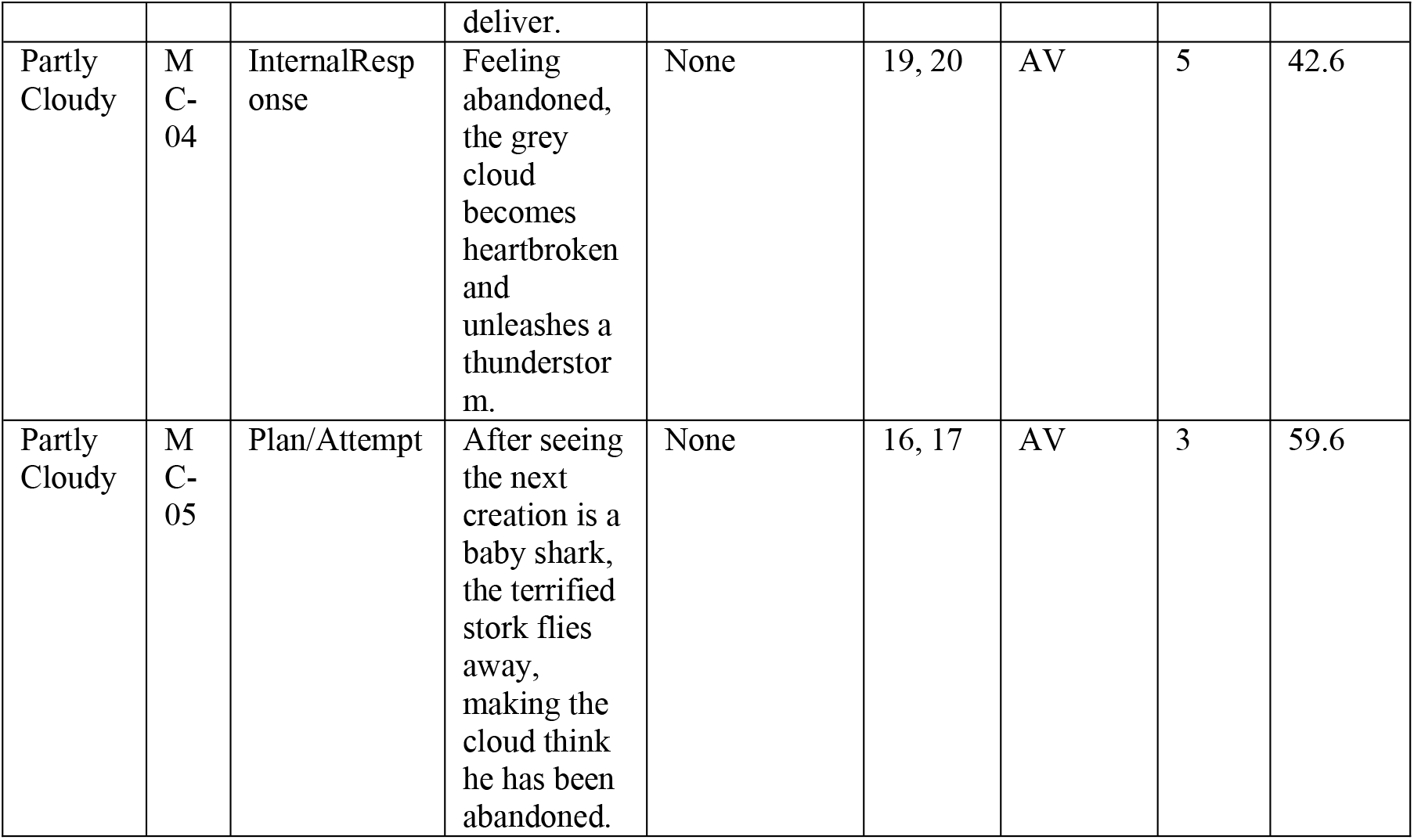
VLM-derived main concept inventories for the eight movie clips, including the movie title, concept narrative role as assigned by the model, concept textual description (used as a reference for scoring participant narrations), subtitle quote(s) and corresponding time stamp(s) (in seconds) cited by the model to support the concept, grounding modality (subtitles - S, audiovisual content - AV, or both), number of VLM runs (out of five) that proposed the concept, and ratio of HC producing the concept (%).

**Table S2.** Clip-wise main concept description, including number of inventory main concepts, mean and range of inventory concepts produced by HC (%), and group difference (PWA versus HC) in main concept completeness and semantic distance from inventory centroid (Cohen’s d).

| Clip | Number of Inventory Concepts | Mean % HC produced | Range % HC produced | d completeness | d distance |
| --- | --- | --- | --- | --- | --- |
| Akeelah and the Bee | 4 | 35.4 | 24.4 to 46.3 | -0.93 | 0.82 |
| Catch Me If You Can | 5 | 62.4 | 40.5 to 88.1 | -1.50 | 0.94 |
| Good Will Hunting | 5 | 32.6 | 23.3 to 44.2 | -0.95 | 1.38 |
| Miracle | 6 | 33.4 | 27.6 to 48.3 | -0.86 | 1.33 |
| Moonlight | 4 | 46.2 | 33.3 to 84.8 | -0.99 | 0.99 |
| No Country for Old Men | 6 | 29.6 | 22.5 to 40.0 | -0.98 | 0.83 |
| The Parent Trap | 4 | 50.6 | 40.9 to 75.0 | -1.19 | 1.00 |
| Partly Cloudy | 5 | 59.2 | 42.6 to 74.5 | -1.10 | 0.98 |

**Table S3.**
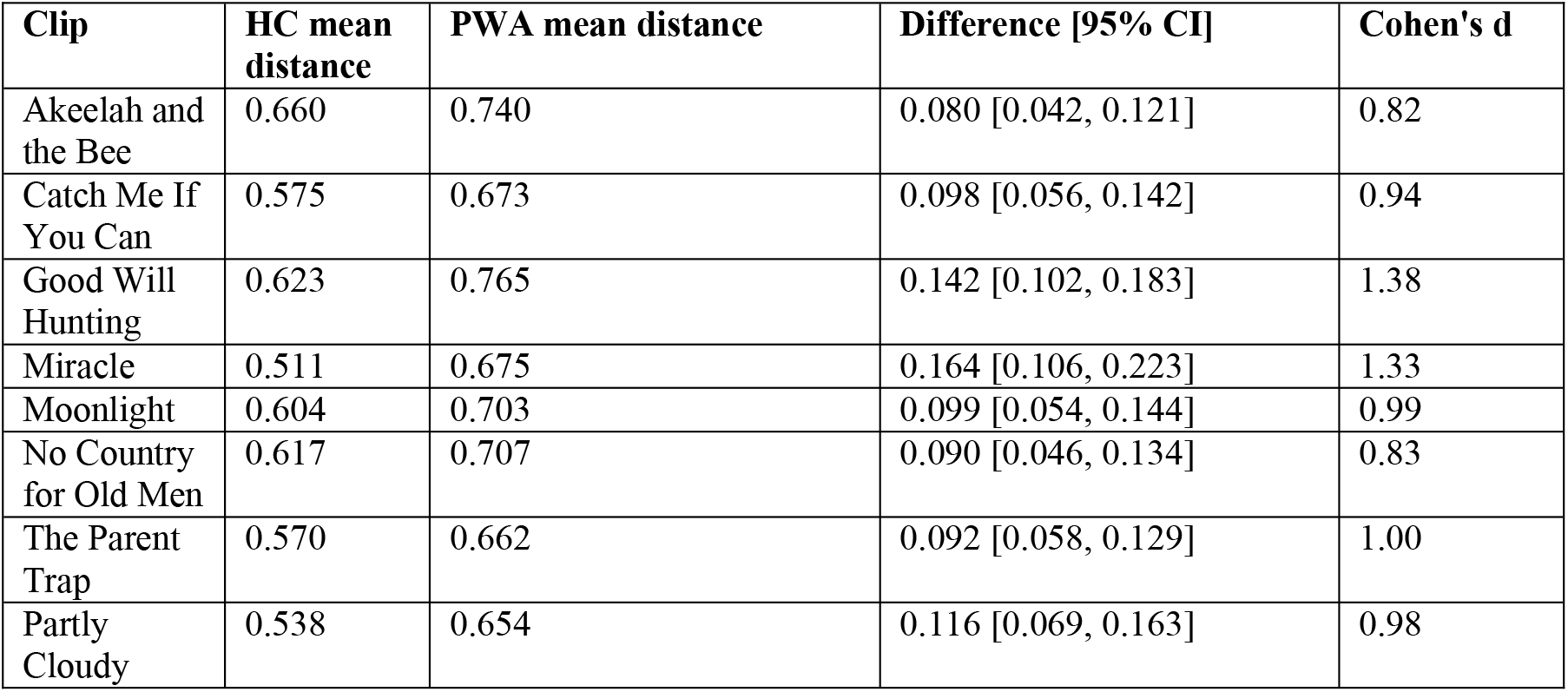
Clip-wise semantic distance to the main concept inventory centroid, including mean cosine distance to the inventory centroid in each group (HC, PWA), and the group difference in semantic distance to the inventory (PWA minus HC) with its 95% confidence interval based on 5,000 bootstrap resamples of participants within group, and Cohen’s d.

| Clip | HC mean distance | PWA mean distance | Difference [95% CI] | Cohen's d |
| --- | --- | --- | --- | --- |
| Akeelah and the Bee | 0.660 | 0.740 | 0.080 [0.042, 0.121] | 0.82 |
| Catch Me If You Can | 0.575 | 0.673 | 0.098 [0.056, 0.142] | 0.94 |
| Good Will Hunting | 0.623 | 0.765 | 0.142 [0.102, 0.183] | 1.38 |
| Miracle | 0.511 | 0.675 | 0.164 [0.106, 0.223] | 1.33 |
| Moonlight | 0.604 | 0.703 | 0.099 [0.054, 0.144] | 0.99 |
| No Country for Old Men | 0.617 | 0.707 | 0.090 [0.046, 0.134] | 0.83 |
| The Parent Trap | 0.570 | 0.662 | 0.092 [0.058, 0.129] | 1.00 |
| Partly Cloudy | 0.538 | 0.654 | 0.116 [0.069, 0.163] | 0.98 |

**Table S4.**
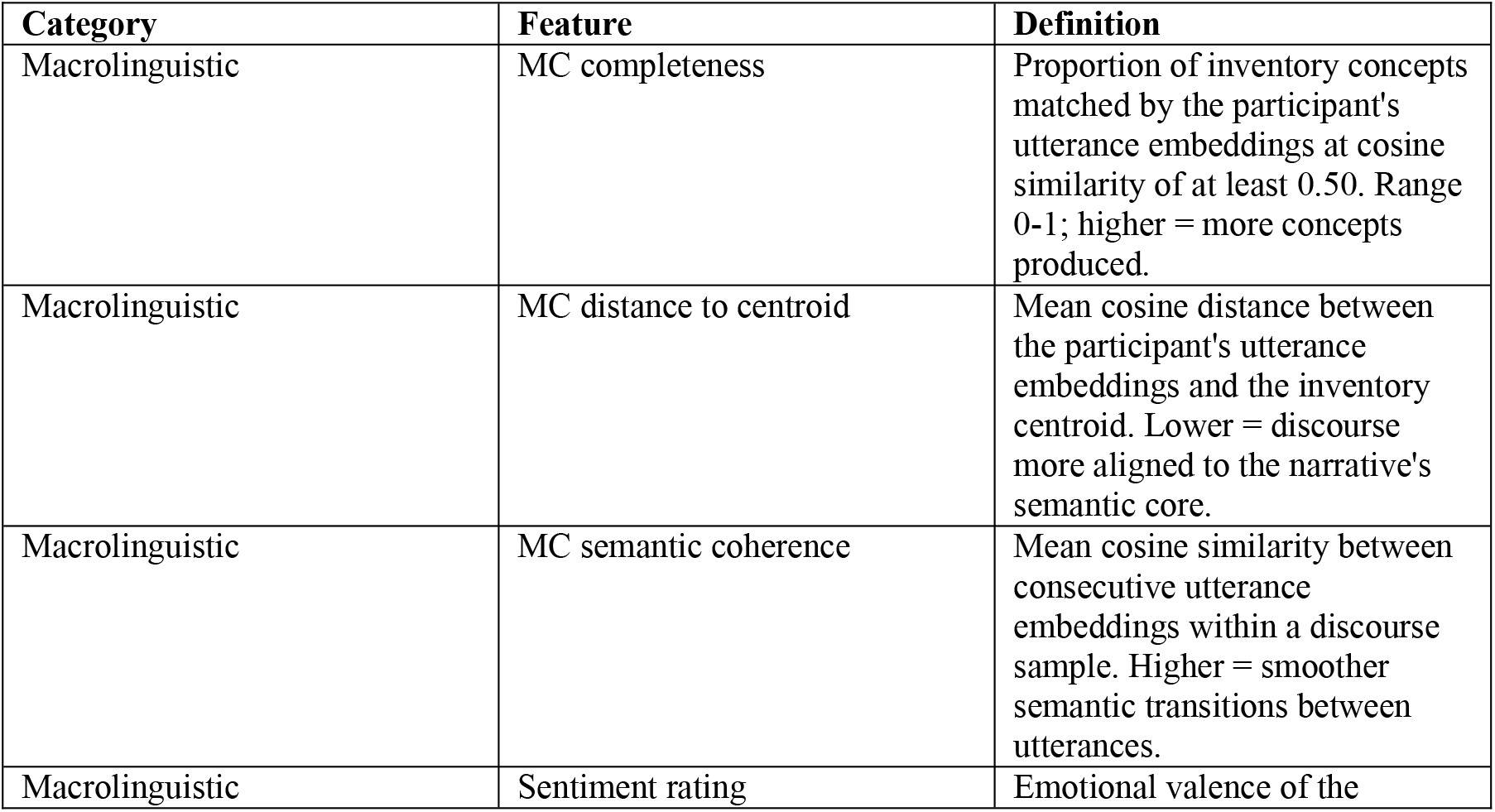
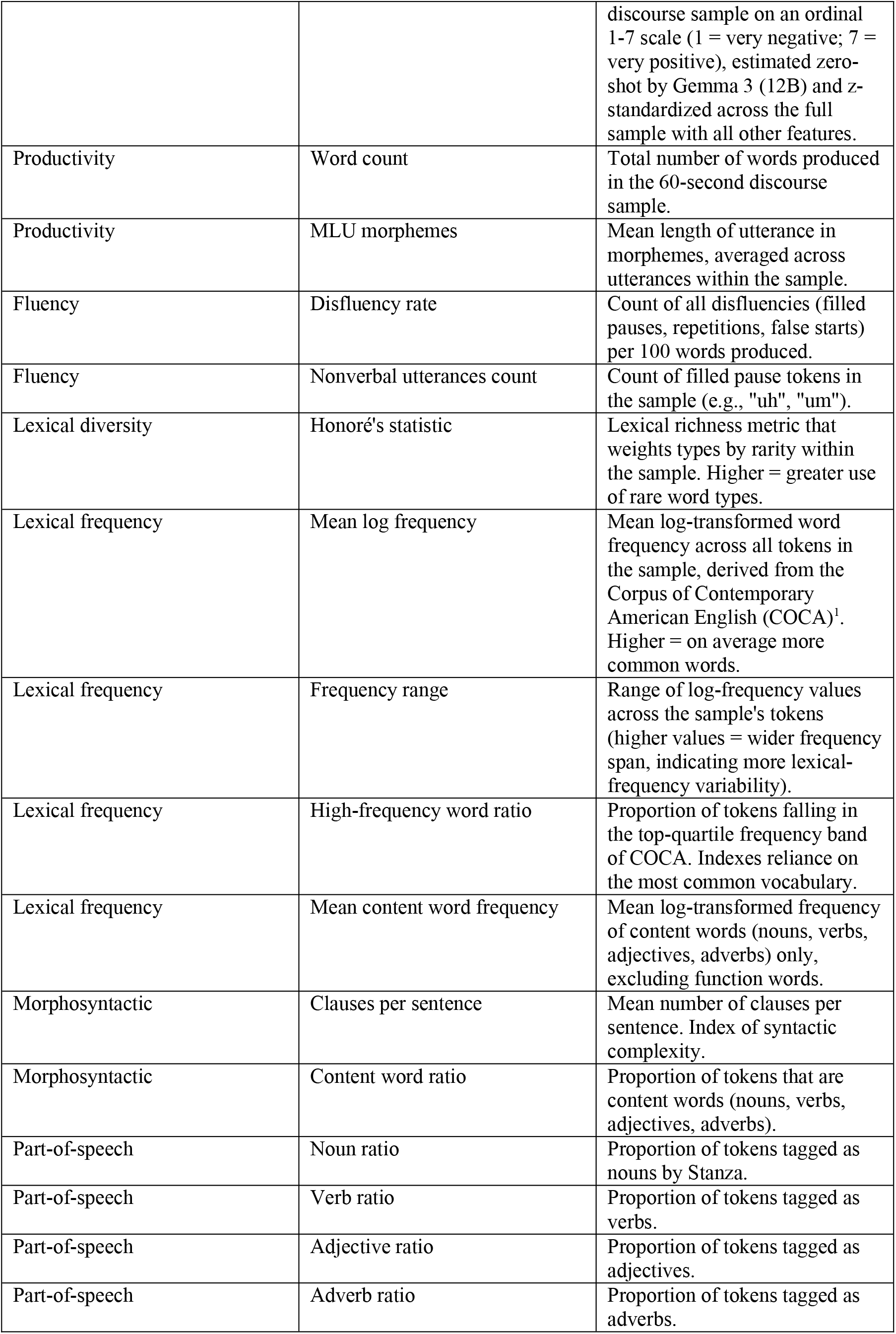
Feature definitions for the 19 primary discourse features. Fourteen further features were removed at the collinearity step because they correlated at |r| > 0.60 with a feature retained here: filled pauses per 100 words, Brunet’s index, function word ratio, mean word frequency, median word frequency, MLU in words, out-of-vocabulary ratio, mean syllable count, type-token ratio, word repetitions, and four main concept variants (coverage normalized, concept efficiency, topic stability, and coherence composite). Three derivative summary statistics (the maximum, minimum and standard deviation of syllable count per utterance) were removed before that step.

**Table S5.** ANCOVA to assess the effect of session modality (in-person, remote) on basic discourse features in PWA (n = 54), adjusted for age and AQ.

| Feature | $\beta$ (session modality) | SE | t | p | N (in-person) | N (remote) |
| --- | --- | --- | --- | --- | --- | --- |
| Word count | -0.217 | 0.201 | -1.078 | 0.286 | 18 | 36 |
| Mean length of utterance | -0.328 | 0.167 | -1.96 | 0.056 | 18 | 36 |

**Table S6.** Per-clip, per-feature effect sizes (PWA minus HC) in standardized z-score units.

| Clip | Feature | Cohen's d (PWA - HC) |
| --- | --- | --- |
| AkeelahAndTheBee | Adjective ratio | +0.06 |
| AkeelahAndTheBee | Adverb ratio | -0.26 |
| AkeelahAndTheBee | Clauses per sentence | +0.25 |
| AkeelahAndTheBee | Content word ratio | -0.30 |
| AkeelahAndTheBee | Disfluency rate | +0.50 |
| AkeelahAndTheBee | Frequency range | -0.66 |
| AkeelahAndTheBee | High-frequency word ratio | +0.42 |
| AkeelahAndTheBee | Honoré's statistic | -0.51 |
| AkeelahAndTheBee | MC distance to centroid | +0.82 |
| AkeelahAndTheBee | MC completeness | -0.92 |
| AkeelahAndTheBee | MC semantic coherence | +0.30 |
| AkeelahAndTheBee | Mean content word frequency | +0.68 |
| AkeelahAndTheBee | Mean log frequency | +0.27 |
| AkeelahAndTheBee | MLU morphemes | -0.16 |
| AkeelahAndTheBee | Noun ratio | -0.30 |
| AkeelahAndTheBee | Sentiment rating | -0.82 |
| AkeelahAndTheBee | Nonverbal utterances count | +0.30 |
| AkeelahAndTheBee | Verb ratio | +0.18 |
| AkeelahAndTheBee | Word count | -0.60 |
| CatchMeIfYouCan | Adjective ratio | -0.37 |
| CatchMeIfYouCan | Adverb ratio | -0.17 |
| CatchMeIfYouCan | Clauses per sentence | -0.08 |
| CatchMeIfYouCan | Content word ratio | -0.06 |
| CatchMeIfYouCan | Disfluency rate | +0.71 |
| CatchMeIfYouCan | Frequency range | -0.60 |
| CatchMeIfYouCan | High-frequency word ratio | +0.26 |
| CatchMeIfYouCan | Honoré's statistic | -0.46 |
| CatchMeIfYouCan | MC distance to centroid | +0.94 |
| CatchMeIfYouCan | MC completeness | -1.50 |
| CatchMeIfYouCan | MC semantic coherence | +0.10 |
| CatchMeIfYouCan | Mean content word frequency | +1.24 |
| CatchMeIfYouCan | Mean log frequency | +0.29 |
| CatchMeIfYouCan | MLU morphemes | -0.54 |
| CatchMeIfYouCan | Noun ratio | -0.00 |
| CatchMeIfYouCan | Sentiment rating | -0.44 |
| CatchMeIfYouCan | Nonverbal utterances count | +0.73 |
| CatchMeIfYouCan | Verb ratio | +0.29 |
| CatchMeIfYouCan | Word count | -0.85 |
| GoodWillHunting | Adjective ratio | +0.20 |
| GoodWillHunting | Adverb ratio | -0.27 |
| GoodWillHunting | Clauses per sentence | -0.18 |
| GoodWillHunting | Content word ratio | -0.38 |
| GoodWillHunting | Disfluency rate | +0.76 |
| GoodWillHunting | Frequency range | -0.60 |
| GoodWillHunting | High-frequency word ratio | +0.15 |
| GoodWillHunting | Honoré's statistic | -0.35 |
| GoodWillHunting | MC distance to centroid | +1.37 |
| GoodWillHunting | MC completeness | -0.95 |
| GoodWillHunting | MC semantic coherence | -0.55 |
| GoodWillHunting | Mean content word frequency | +1.04 |
| GoodWillHunting | Mean log frequency | +0.44 |
| GoodWillHunting | MLU morphemes | -0.52 |
| GoodWillHunting | Noun ratio | -0.30 |
| GoodWillHunting | Sentiment rating | +0.49 |
| GoodWillHunting | Nonverbal utterances count | +0.22 |
| GoodWillHunting | Verb ratio | +0.04 |
| GoodWillHunting | Word count | -0.78 |
| Miracle | Adjective ratio | +0.17 |
| Miracle | Adverb ratio | +0.28 |
| Miracle | Clauses per sentence | +0.37 |
| Miracle | Content word ratio | -0.43 |
| Miracle | Disfluency rate | +0.79 |
| Miracle | Frequency range | -0.32 |
| Miracle | High-frequency word ratio | +0.35 |
| Miracle | Honoré's statistic | -0.64 |
| Miracle | MC distance to centroid | +1.33 |
| Miracle | MC completeness | -0.86 |
| Miracle | MC semantic coherence | -0.28 |
| Miracle | Mean content word frequency | +0.08 |
| Miracle | Mean log frequency | +0.54 |
| Miracle | MLU morphemes | -0.10 |
| Miracle | Noun ratio | -0.52 |
| Miracle | Sentiment rating | -0.63 |
| Miracle | Nonverbal utterances count | +0.74 |
| Miracle | Verb ratio | -0.07 |
| Miracle | Word count | +0.02 |
| Moonlight | Adjective ratio | -0.11 |
| Moonlight | Adverb ratio | +0.42 |
| Moonlight | Clauses per sentence | -0.02 |
| Moonlight | Content word ratio | +0.04 |
| Moonlight | Disfluency rate | +0.91 |
| Moonlight | Frequency range | -0.60 |
| Moonlight | High-frequency word ratio | -0.16 |
| Moonlight | Honoré's statistic | -0.60 |
| Moonlight | MC distance to centroid | +0.99 |
| Moonlight | MC completeness | -0.99 |
| Moonlight | MC semantic coherence | -0.22 |
| Moonlight | Mean content word frequency | +0.64 |
| Moonlight | Mean log frequency | +0.29 |
| Moonlight | MLU morphemes | -0.74 |
| Moonlight | Noun ratio | -0.23 |
| Moonlight | Sentiment rating | -1.07 |
| Moonlight | Nonverbal utterances count | +1.00 |
| Moonlight | Verb ratio | +0.23 |
| Moonlight | Word count | -0.45 |
| NoCountryForOldMen | Adjective ratio | -0.09 |
| NoCountryForOldMen | Adverb ratio | -0.41 |
| NoCountryForOldMen | Clauses per sentence | -0.26 |
| NoCountryForOldMen | Content word ratio | -0.29 |
| NoCountryForOldMen | Disfluency rate | +0.57 |
| NoCountryForOldMen | Frequency range | -0.61 |
| NoCountryForOldMen | High-frequency word ratio | -0.02 |
| NoCountryForOldMen | Honoré's statistic | -0.30 |
| NoCountryForOldMen | MC distance to centroid | +0.83 |
| NoCountryForOldMen | MC completeness | -0.98 |
| NoCountryForOldMen | MC semantic coherence | -0.13 |
| NoCountryForOldMen | Mean content word frequency | +0.57 |
| NoCountryForOldMen | Mean log frequency | +0.19 |
| NoCountryForOldMen | MLU morphemes | -0.54 |
| NoCountryForOldMen | Noun ratio | -0.13 |
| NoCountryForOldMen | Sentiment rating | -0.39 |
| NoCountryForOldMen | Nonverbal utterances count | +0.65 |
| NoCountryForOldMen | Verb ratio | +0.16 |
| NoCountryForOldMen | Word count | -0.78 |
| ParentTrap | Adjective ratio | -0.54 |
| ParentTrap | Adverb ratio | -0.20 |
| ParentTrap | Clauses per sentence | -0.08 |
| ParentTrap | Content word ratio | -0.34 |
| ParentTrap | Disfluency rate | +0.86 |
| ParentTrap | Frequency range | -0.59 |
| ParentTrap | High-frequency word ratio | +0.53 |
| ParentTrap | Honoré's statistic | -0.57 |
| ParentTrap | MC distance to centroid | +1.00 |
| ParentTrap | MC completeness | -1.19 |
| ParentTrap | MC semantic coherence | +0.25 |
| ParentTrap | Mean content word frequency | +0.09 |
| ParentTrap | Mean log frequency | +0.60 |
| ParentTrap | MLU morphemes | -0.62 |
| ParentTrap | Noun ratio | -0.08 |
| ParentTrap | Sentiment rating | -0.34 |
| ParentTrap | Nonverbal utterances count | +0.55 |
| ParentTrap | Verb ratio | +0.03 |
| ParentTrap | Word count | -0.94 |
| PartlyCloudy | Adjective ratio | -0.18 |
| PartlyCloudy | Adverb ratio | -0.27 |
| PartlyCloudy | Clauses per sentence | -0.13 |
| PartlyCloudy | Content word ratio | -0.67 |
| PartlyCloudy | Disfluency rate | +0.88 |
| PartlyCloudy | Frequency range | -0.51 |
| PartlyCloudy | High-frequency word ratio | +0.30 |
| PartlyCloudy | Honoré's statistic | -0.44 |
| PartlyCloudy | MC distance to centroid | +0.98 |
| PartlyCloudy | MC completeness | -1.10 |
| PartlyCloudy | MC semantic coherence | -0.02 |
| PartlyCloudy | Mean content word frequency | +0.79 |
| PartlyCloudy | Mean log frequency | +0.41 |
| PartlyCloudy | MLU morphemes | -0.47 |
| PartlyCloudy | Noun ratio | -0.39 |
| PartlyCloudy | Sentiment rating | -0.72 |
| PartlyCloudy | Nonverbal utterances count | +0.77 |
| PartlyCloudy | Verb ratio | -0.35 |
| PartlyCloudy | Word count | -1.38 |

**Table S7.** Sensitivity of the main concept measures to the utterance-to-concept matching threshold. An utterance credits its nearest inventory concept when cosine similarity exceeds the threshold; 0.50 was used throughout the main analyses and was set a priori rather than calibrated on these data. Completeness is the mean percentage of a clip’s inventory concepts produced, across all participants. Cohen’s d and the partial correlation with the WAB-R Aphasia Quotient (AQ, range 0–100) are computed at the participant level as in Tables 1 and 2. AUC and accuracy are from the same LASSO model with leave-two-out cross-validation. All group differences were significant at p < .001.

| Matching threshold | Mean completeness (%) | Cohen's d (PWA – HC) | r with AQ | AUC | Accuracy |
| --- | --- | --- | --- | --- | --- |
| 0.40 | 47.5 | –1.57 | 0.74 | 0.939 | 0.895 |
| 0.50 (used here) | 31.3 | –1.84 | 0.66 | 0.939 | 0.905 |
| 0.60 | 13.8 | –1.81 | 0.53 | 0.940 | 0.895 |
| 0.70 | 2.8 | –1.12 | 0.36 | 0.944 | 0.895 |

**Table S8.** Sensitivity analyses for the group comparisons. For each of the 19 features: the ratio of group standard deviations; Cohen’s d computed from raw group means and from the age-adjusted ANCOVA contrast; the ANCOVA p-value with ordinary and with heteroskedasticity-consistent (HC3) standard errors (both FDR-corrected); d after residualizing the feature on word count and age; and d after standardizing each feature within clip before aggregating to the participant.

| Feature | SD ratio | d (raw) | d (age-adj.) | q (OLS) | q (HC3) | d (length-adj.) | d (within-clip) |
| --- | --- | --- | --- | --- | --- | --- | --- |
| Nonverbal utterances count | 1.74 | 0.62 | 0.54 | 0.025 | 0.013 | 0.51 | 0.64 |
| Word count | 0.88 | -0.84 | -1.07 | <.001 | <.001 | — | -0.83 |
| Adjective ratio | 1.66 | -0.22 | -0.21 | 0.367 | 0.382 | -0.11 | -0.22 |
| Adverb ratio | 1.35 | -0.22 | -0.30 | 0.201 | 0.188 | 0.11 | -0.21 |
| Clauses per sentence | 1.75 | -0.09 | -0.09 | 0.736 | 0.721 | -0.07 | -0.09 |
| Content word ratio | 2.90 | -0.45 | -0.49 | 0.039 | 0.038 | -0.39 | -0.44 |
| Disfluency rate | 5.60 | 1.07 | 1.15 | <.001 | <.001 | 0.68 | 1.19 |
| Frequency range | 2.64 | -0.78 | -0.81 | <.001 | <.001 | -0.19 | -0.86 |
| High-frequency word ratio | 1.49 | 0.44 | 0.41 | 0.088 | 0.071 | 0.63 | 0.53 |
| Honoré's statistic | 1.36 | -0.84 | -0.99 | <.001 | <.001 | -0.73 | -0.82 |
| Mean content word frequency | 4.27 | 0.37 | 0.31 | 0.196 | 0.218 | 0.47 | 0.70 |
| Mean log frequency | 2.42 | 0.41 | 0.41 | 0.089 | 0.071 | 0.68 | 0.44 |
| MLU morphemes | 1.07 | -0.76 | -0.72 | 0.003 | 0.002 | -0.40 | -0.79 |
| Noun ratio | 1.68 | -0.36 | -0.33 | 0.172 | 0.112 | -0.58 | -0.39 |
| Verb ratio | 2.12 | 0.03 | -0.04 | 0.881 | 0.864 | 0.16 | 0.03 |
| Sentiment rating | 1.73 | -0.87 | -0.70 | 0.003 | 0.002 | -0.31 | -0.85 |
| MC completeness | 1.09 | -1.84 | -1.88 | <.001 | <.001 | -1.01 | -1.83 |
| MC semantic coherence | 2.56 | -0.07 | -0.03 | 0.881 | 0.864 | -0.38 | -0.04 |
| MC distance to centroid | 1.79 | 1.41 | 1.28 | <.001 | <.001 | 0.87 | 1.39 |

**Table S9.** Classification model comparison. Each model was fitted with the leave-two-out cross-validation of the primary analysis (2,754 folds; λ selected by nested 5-fold cross-validation within each fold). AUCs are given with 95% bootstrap confidence intervals (2,000 resamples); nested models were compared with DeLong’s test for correlated ROC curves. The full model exceeded the microlinguistic-only model by ΔAUC = 0.034 (95% CI [−0.003, 0.070]) and the macrolinguistic-only model by ΔAUC = 0.039 (95% CI [−0.025, 0.104]).

| Model | Features | AUC | 95% CI |
| --- | --- | --- | --- |
| micro only | 15 | 0.914 | [0.850, 0.968] |
| macro only | 4 | 0.908 | [0.843, 0.966] |
| micro + macro | 19 | 0.948 | [0.897, 0.986] |
| age only | 1 | 0.671 | [0.571, 0.768] |
| micro + macro + age (unpenalized) | 20 | 0.962 | [0.916, 0.993] |
| DeLong: full vs microlinguistic only | — | Z = 1.81 | p = 0.070 |
| DeLong: full vs macrolinguistic only | — | Z = 1.19 | p = 0.235 |

**Table S10.** Hierarchical regression predicting aphasia severity (WAB-R AQ) in the PWA subsample (n = 54). The microlinguistic block comprises the nine microlinguistic features significantly associated with AQ in Table 2. The final row enters the two main concept features in the reverse order.

| Step | Predictors | R<sup>2</sup> | Adjusted R<sup>2</sup> | $\Delta R^2$ | F change | df | p |
| --- | --- | --- | --- | --- | --- | --- | --- |
| 1. age | 1 | 0.001 | -0.019 | — | — | — | — |
| 2. + microlinguistic | 10 | 0.723 | 0.658 | 0.722 | 12.43 | 9, 43 | <.001 |
| 3. + MC completeness | 11 | 0.761 | 0.699 | 0.039 | 6.81 | 1, 42 | 0.013 |
| 4. + MC distance to centroid | 12 | 0.830 | 0.780 | 0.069 | 16.62 | 1, 41 | <.001 |
| Reverse order | completeness entered after distance | — | — | 0.003 | — | 1, 41 | 0.417 |

**Table S11.** Bootstrap stability of the classification coefficients. The final model was refitted on 2,000 bootstrap resamples of participants at the selected penalty. Columns give the coefficient in the reported model, the 2.5th to 97.5th percentile interval and the bootstrap standard error across resamples, the percentage of resamples in which the coefficient reversed sign, and the percentage in which it was zero.

| Feature | Coefficient | 95% bootstrap CI | Bootstrap SE | Sign flips (%) | Zero in resample (%) |
| --- | --- | --- | --- | --- | --- |
| MC distance to centroid | 2.538 | [0.00, 3.83] | 1.088 | 0.0 | 9.9 |
| Mean log frequency | 2.231 | [0.00, 3.25] | 1.032 | 0.0 | 17.7 |
| MC completeness | -1.877 | [-4.62, -0.35] | 1.113 | 0.0 | 0.8 |
| Disfluency rate | 1.640 | [0.00, 3.88] | 1.040 | 0.0 | 7.8 |
| Nonverbal utterances count | 1.611 | [0.00, 3.55] | 0.861 | 0.1 | 2.5 |
| Word count | -1.385 | [-2.54, 0.00] | 0.768 | 0.0 | 14.2 |
| Clauses per sentence | -1.042 | [-2.66, 0.00] | 0.812 | 0.1 | 31.2 |
| Adjective ratio | -0.816 | [-2.45, 0.00] | 0.710 | 0.2 | 12.2 |
| Adverb ratio | -0.543 | [-1.85, 0.00] | 0.561 | 0.2 | 26.1 |
| Honoré's statistic | -0.525 | [-2.26, 0.00] | 0.659 | 0.0 | 23.9 |
| Sentiment rating | -0.414 | [-2.44, 0.00] | 0.710 | 1.0 | 45.7 |
| MLU morphemes | -0.251 | [-2.63, 0.00] | 0.781 | 0.1 | 22.9 |
| Mean content word frequency | 0.024 | [0.00, 5.99] | 1.957 | 0.0 | 54.1 |
| Content word ratio | 0.000 | [-0.83, 0.00] | 0.238 | — | 92.2 |
| Frequency range | 0.000 | [-0.61, 0.00] | 0.209 | — | 90.5 |
| High-frequency word ratio | 0.000 | [0.00, 2.00] | 0.598 | — | 66.1 |
| Noun ratio | 0.000 | [0.00, 1.35] | 0.364 | — | 72.7 |
| Verb ratio | 0.000 | [-0.71, 0.26] | 0.217 | — | 81.0 |
| MC semantic coherence | 0.000 | [-1.54, 0.00] | 0.456 | — | 74.0 |

**Table S12.**
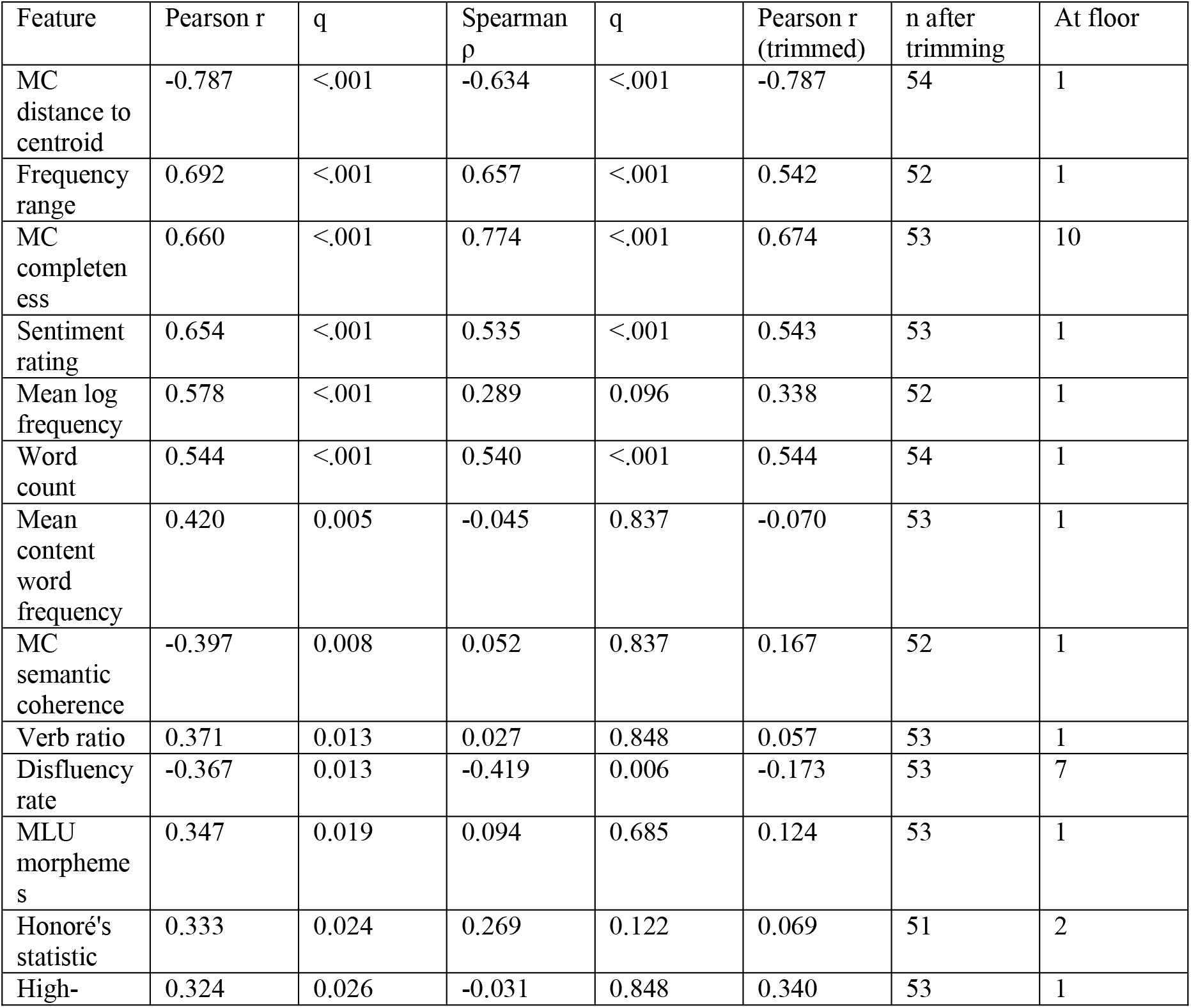
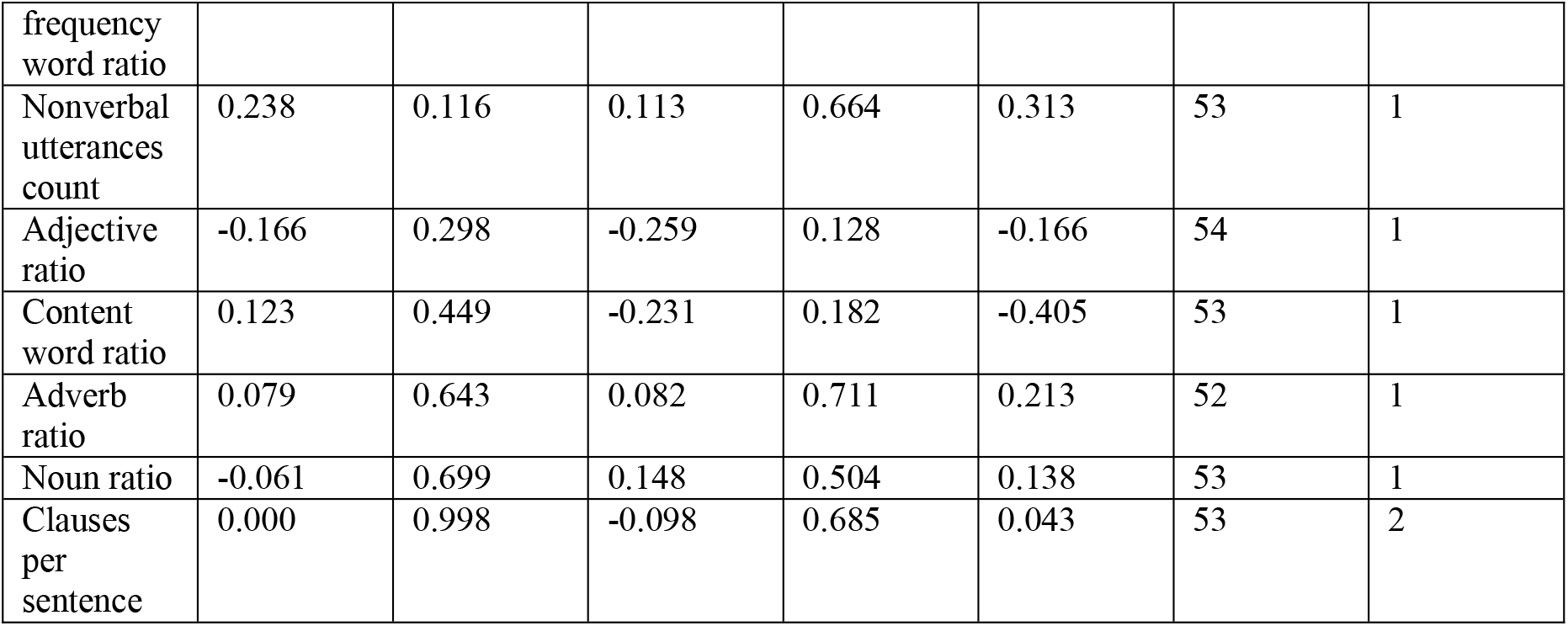
Partial correlations with aphasia severity, by estimator. Pearson and Spearman partial correlations with the WAB-R AQ, adjusting for age, with FDR-corrected p-values; and the Pearson correlation recomputed after removing observations beyond |z| = 2.5 on the feature. The last two columns give the number of participants retained after trimming and the number at the feature’s minimum. The six features significant under both estimators are those shown in main-text Figure 3.

**Table S13.**
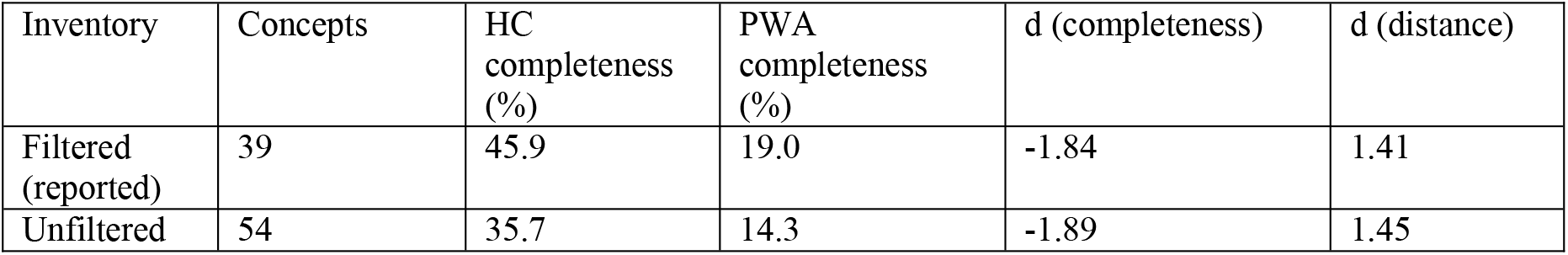
Sensitivity of the main concept measures to the healthy-control production criterion. Concepts produced by fewer than 20% of controls were removed from the panel of 54, leaving the 39 reported inventory concepts; all participants were also scored against the unfiltered 54-concept inventory. Removing any single control from the production criterion leaves the inventory unchanged (no concept’s retention status changes under leave-one-control-out; 29 to 47 controls per clip).

| Inventory | Concepts | HC completeness (%) | PWA completeness (%) | d (completeness) | d (distance) |
| --- | --- | --- | --- | --- | --- |
| Filtered (reported) | 39 | 45.9 | 19.0 | -1.84 | 1.41 |
| Unfiltered | 54 | 35.7 | 14.3 | -1.89 | 1.45 |

**Figure S1.**
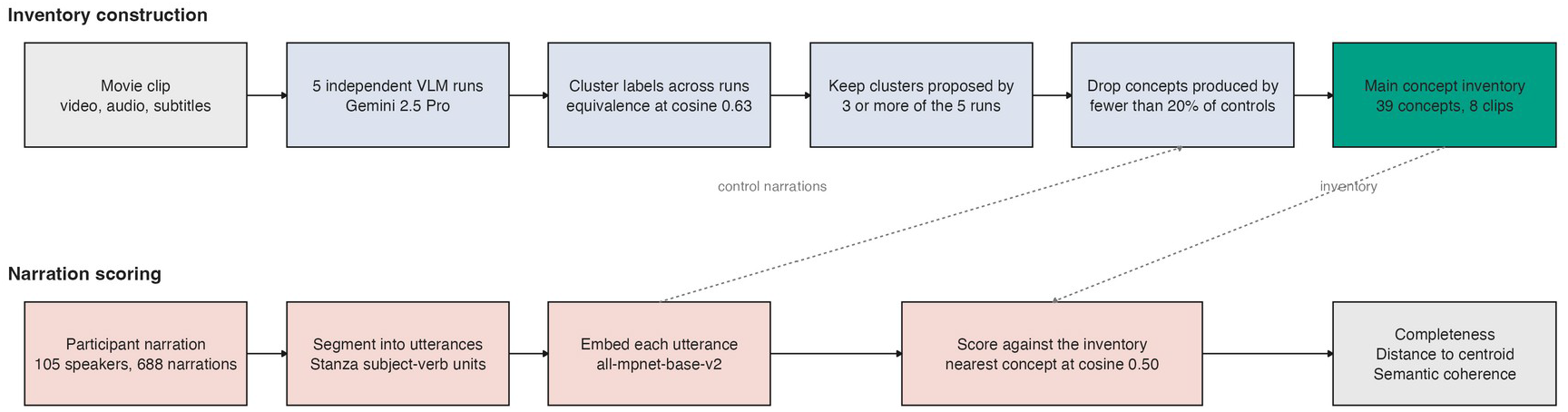
The analysis pipeline. The upper lane builds a main concept inventory from the clip: five independent vision-language model runs, clustering of concept labels across runs at the calibrated equivalence threshold, retention of clusters proposed by at least three of the five runs, and removal of concepts produced by fewer than 20% of healthy controls. The lower lane scores participant narrations against that inventory. Dashed arrows mark the two points at which the lanes meet: control narrations set the 20% production floor, and the finished inventory is the reference against which all narrations are scored.

